# A deep-learning based biomarker of systemic cellular senescence burden to predict mortality and health outcomes

**DOI:** 10.64898/2026.03.20.26348913

**Authors:** Shangshu Zhao, Chia-Ling Kuo, Eric J. Lenze, Julie L. Wetherell, Laura Haynes, Perla El-Ahmad, Richard Fortinsky, George Kuchel, Trevor Harris, Breno S. Diniz

## Abstract

**Introduction:** The accumulation of senescent cells is a recognized hallmark of biological aging and is associated with the onset of multiple chronic medical conditions. Senescent cells exhibit a distinct secretory profile, known as the senescence-associated secretory phenotype (SASP), which can propagate cellular senescence to neighboring and distant tissues. Measuring SASP factors in blood serves as a practical proxy for cellular senescence burden and may help track disease states and intervention outcomes.

**Methods:** We developed and validated a composite SASP Score by integrating large-scale population proteomics data with a semi-supervised deep learning framework. The analytical workflow included: (1) selection of biologically curated SASP proteins; (2) development of a Guided autoencoder with Transformer (GAET) model using data from the UK Biobank Pharma Proteomics Project (UKB-PPP); (3) internal evaluation and association analyses within the UK Biobank; and (4) external validation and longitudinal assessment in an independent randomized clinical trial cohort.

**Results:** The deep learning-based SASP Score was a strong, independent predictor of mortality risk and incident serious, chronic medical conditions (e.g., dementia, COPD, myocardial infarction, stroke). In an independent cohort, multimodal exercise significantly changed the SASP Score trajectory over 18 months.

**Discussion:** Our findings support the potential of a deep learning-derived SASP Score as a biomarker for systemic cellular senescence burden. Our statistical approach can offer enhanced interpretability and cross-platform utility, providing a valuable tool for aging research and the evaluation of geroscience-guided interventions.

## Introduction

Cellular senescence is one of the main hallmarks of biological aging^1^. It is characterized by a state of permanent cell cycle arrest, with upregulation of cyclin-dependent kinase 2 inhibitors (e.g., p16 and p21), upregulation of anti-apoptotic factors, nuclear changes (e.g., increased senescence-associated heterochromatic foci (SAHF), telomere-associated foci (TAF) formation, and loss of lamin B1 (LMNB1), cellular morphological changes (e.g., cell enlargement, vacuolization), among others^2,3^. Although senescent cells accumulate in multiple tissues with aging, they constitute a very small proportion of the total cells in a given tissue, making their identification challenging in human studies^3^.

Despite being in permanent cell-cycle arrest, senescent cells remain metabolically active and exhibit a gradual change in their secretome profile, known as the senescence-associated secretory phenotype (SASP)^4^. The SASP can amplify the impact of cell-intrinsic proliferative arrest in neighboring and distant tissues, leading to the accumulation of senescent cells, thereby contributing to the emergence of multiple chronic, age-related diseases and increased mortality risk in older adults^5,6^. Therefore, the accumulation of SASP factors serves as a proxy for increased cellular senescence burden, and, given that they can be easily evaluated in a blood draw, they are useful for tracking cellular senescence burden across different disease states and the impact of intervention^7^.

Recent work has demonstrated that SASP factors are independent predictors of age-related diseases and mortality^8,9^, including composite biomarker indices based on SASP factors that have been associated with various neuropsychiatric disorders^10–13^. Importantly, despite recent advances, analyzing SASP factors individually may not fully reflect SASP complexity and can limit the interpretability of results. Moreover, composite biomarker indices developed using linear dimensionality-reduction techniques (e.g., principal component analysis) may not capture complex nonlinear relationships among these factors. In this study, we developed the SASP Score based on multiple SASP proteins using a semi-supervised deep learning model. This model addresses two major limitations noted previously: first, it creates a single SASP composite index and accounts for complex, non-linear relationships among SASP proteins, and second, it enables its robust application across multiple proteomic platforms without the need for special data transformation.

## Methods

We developed and validated the SASP Score by integrating large-scale population proteomics with a semi-supervised deep learning framework. The overall analytical workflow comprised four key components: (1) selection of biologically curated SASP proteins; (2) development of a Guided AutoEncoder with Transformer (GAET) model using UK Biobank Pharma Proteomics Project (UKB-PPP) data; (3) internal evaluation and association analyses within the UK Biobank; and (4) external validation and longitudinal assessment in an independent randomized clinical trial cohort. Details of the study populations, proteomic measurements, model development, post-development adaptation, and statistical analyses are described below.

### UK Biobank cohort

The UK Biobank is a prospective population-based cohort in the United Kingdom^14^. Over 500,000 participants aged 40 to 70 were recruited between 2006 and 2010. At baseline, participants provided sociodemographic, lifestyle, environmental, clinical, and physical assessment data, and biological samples were collected for future assays. Participants have been followed longitudinally through linkage with electronic health records for disease diagnoses and mortality^15^.

### UK Biobank Pharma Proteomics Project data

Participants who supplied blood samples at baseline were selected for inclusion in the UK Biobank Pharma Proteomics Project (UKB-PPP) (n=54,219)^16^. Most of the included participants were randomly selected from the UK Biobank baseline cohort (n=46,595, 85.9%). Additional participants were selected by the UKB-PPP consortium of 13 biopharmaceutical companies based on specific diseases or ancestry of interest (n = 6,376), and from the COVID-19 repeat imaging study (n = 1,268). Expression levels of 2,923 plasma proteins were measured using the Olink Explore 3072 assay (Olink, Sweden) and normalized using a two-step process: within-batch and across-batch intensity normalization. Normalized protein expression (NPX) data were used throughout the present analysis. Proteomic data from 50,997 active UKB-PPP participants were included in the current analysis. Three proteins were excluded due to high missing rates: GLIPR1 (99.7%), NPM1 (74.0%), and PCOLCE (63.6%). Missing values were imputed using the *k*-nearest neighbors (KNN) approach (*k* = 10) with the R package “multiUS” (https://cran.r-project.org/web/packages/multiUS/index.html).

### Selection of the SASP proteins

After an initial search across multiple resources (*e.g*., CellAge^17^, SenNet Consortium^18,19^, SASP Atlas^20^), prior work from our group^21^, and manual curation of the literature, we identified a common set of 38 plasma proteins available in the UKB-PPP that are consistently dysregulated across different cell types after exposures to various senescence inducers. To be included in this panel, protein expression changes should be in the same direction across senescence-inducing experiments for different cell types, when this information was available. Based on these criteria, a final set of 38 proteins was selected (gene symbols: ANG, CCL2, CCL3, CCL4, CCL13, CCL20, CHI3L1, CSF2, CXCL1, CXCL8, CXCL9, CXCL10, FGA, FGF2, FSTL3, GDF15, HGF, ICAM1, IGFBP2, IGFBP6, IL1B, IL4, IL5, IL6, IL6ST, LEP, MIF, MMP12, PGF, PLAUR, SERPIN1, TF, TIMP1, TNFRSF1A, TNFRSF1B, TNFRSF11B, TNFSF10, VEGFA).

### Development of the SASP Score

Data from the UKB-PPP were randomly split into a training set (85%, n = 43,358) for the SASP Score development and a testing set (15%, n = 7,639) for validation. We developed a composite biomarker score, the SASP Score, based on the selected SASP proteins. Specifically, we developed the Guided AutoEncoder with Transformer (GAET)^22^, a Transformer empowered neural network to learn the SASP Score as the nonlinear latent representation of proteomics data guided by age.This design enables applications across cohorts and protein assays (**Fig. 1**).

**Fig. 1.**
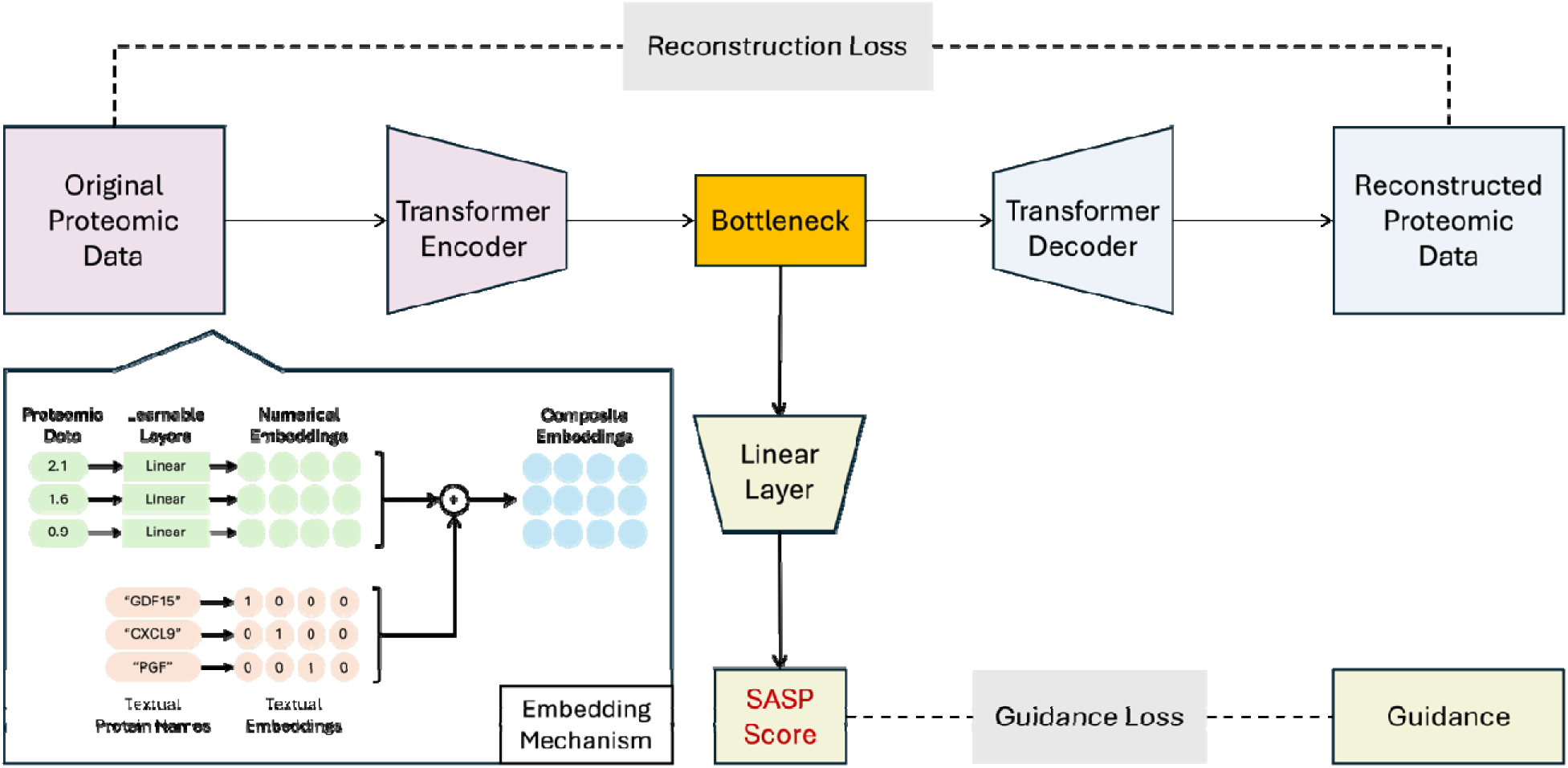
Diagram of Guided AutoEncoder with Transformer (GAET). The GAET model contains three main parts: 1) the encoder to compress input data to a lower dimension (colored in pink), 2) the decoder to reconstruct input data from the compressed representation (colored in light blue), and 3) A linear layer to extract a scalar score from the compressed latent representation (colored in light yellow). A composite embedding mechanism is implemented to encode the textual information of protein names.

The GAET is an extension of classic autoencoders, inheriting its strong capability for nonlinear dimensionality reduction while incorporating an auxiliary guidance head to integrate biologically relevant information. This guidance head provides age-based supervision during training by predicting chronological age from the latent representation, thereby encouraging the model to learn aging-related protein structure while avoiding degenerate or biologically uninformative encodings. Importantly, chronological age is used only to guide model training and is not included in the calculation of the SASP Score once the model is trained or later fine-tuned.

By incorporating the Transformer as the core for both encoder and decoder, GAET is designed to capture complex inter-protein dependencies and facilitate robust translation across cohorts and assays. Through the Transformer, we introduced a dual-embedding mechanism that jointly encodes quantitative protein measurements and textual protein identity, enabling the model to retain protein-level semantics during training and model replication across other cohorts. Technical details of the GAET model are provided in the **Supplementary Methods**.

### External validation cohort: MEDEX

The Mindfulness, Education, and Exercise (MEDEX) study was a randomized clinical trial comparing mindfulness-based stress reduction (MBSR) and multimodal exercise, alone or in combination, to improve cognitive performance in older adults with subjective cognitive complaints (clinicaltrials.gov NCT02665481)^23^. A total of 585 participants aged 65-84 years were recruited and assessed at 3 time points (0, 6, and 18 months) for cognitive (primary) and other health outcomes (secondary). Biological samples collected at each study visit provided longitudinal proteomic data, enabling calculation of the SASP Score at 0, 6, and 18.

The SASP proteins were measured using customized immunoassays from Bio-Techne (Minneapolis, MN) according to the manufacturer’s instructions, using a LUMINEX instrument (Austin, TX). All the samples were run with assays from the same manufacturing lot. The samples for each participant, collected at different assessment time points, were assayed on the same experimental plate. These steps were taken to minimize experimental variation across samples. All assay coefficients of variation were below 18% and were considered acceptable.

The proteomic preprocessing procedures included a normalization step to improve the comparability between samples by reducing the experimental variances and the imputation of missing values. The normalization was performed using the Cycloess method implemented in the R package NormalyzerDE^24^, where Cycloess estimates and removes the bias function of log expression ratio versus log expression abundance (which should be independent). Then the missing values are imputed using multiple imputation (m = 5) and predictive mean matching (method = ‘pmm’) in the R package “mice”^25^.

We applied the GAET model developed in the UKB-PPP to proteomic data measured in the MEDEX cohort to calculate the SASP Score. To adapt the model to this independent cohort, we fine-tuned the pre-trained model using MEDEX data. Chronological age was used as a guidance variable during this process. Fine-tuning allows the model to adjust to differences in measurement platforms and data distributions between cohorts while retaining information learned from the UK Biobank. To reduce overfitting, fine-tuning was performed gradually with conservative tuning settings and stopping criteria (detailed in **Supplementary Methods**). All analyses were conducted using fixed random seeds to ensure reproducibility.

### Statistical analyses

Spearman correlation coefficients were calculated between the SASP Score and chronological age, biological aging measures —including telomere length, proteomic aging clock (PAC)^31^, healthspan proteomic score (HPS)^32^, biological age (BioAge)^23^, phenotypic age (PhenoAge)^22^ — as well as other baseline aging traits including a 49-item frailty index^26^ in the UK Biobank test sample. Details on the development of these biological aging measures are provided in the cited references, and the field IDs used to extract the relevant data are listed in **Table S1.**

The distribution of the SASP Score was compared across subgroups based on a common disease risk factor, including age group, sex, frailty status, waist-to-hip ratio group, and smoking status. Group differences were assessed using Wilcoxon rank-sum tests, with p-values adjusted for multiple testing using the Benjamini-Hochberg false discovery rate (FDR) method^28^. Additionally, the SASP Score distributions in subgroups were visualized using violin plots.

Cox proportional hazards models were used to investigate the associations of the standardized SASP Score with mortality and multiple aging-related diseases during follow-up (censored at death, or the last follow-up date of hospital inpatient data, whichever occurred first), with adjustments for baseline covariates. Mortality was ascertained through linkage to national death registries, and disease outcomes were derived using the UKB first occurrence data, which integrate multi-source data based on ICD-10 codes. Baseline covariates—including age, sex, ethnicity, education level, Townsend deprivation index (higher values associated more material deprivation), smoking status (never, previous, current smoker), alcohol drinker status (never, previous, current drinker), systolic blood pressure, and BMI—were collected from UK Biobank online questionnaires or physical measurements at the baseline assessment. UK Biobank field IDs used to extract covariates and outcomes are provided in **Table S1**. Hazard ratios for each condition per SD increase in the SASP Score were reported with the multiple testing adjusted p-values using the FDR method. Model discrimination was evaluated using Harrell’s C-index, and the 95% confidence intervals were computed using a normal approximation based on the standard error of the C estimate.

Shapley Additive exPlanations (SHAP) analysis^27^ was implemented to evaluate the relative contribution of individual proteins to the SASP Score. For each individual sample, SHAP calculates a contribution of each feature towards the predicted outcome, SASP Score. To compute these contributions, SHAP compares each sample in the test set to the training set representing typical feature levels and estimates how the predicted outcome changes when each feature varies relative to the reference set. Feature importance was then summarized and ranked globally by taking the mean absolute SHAP value across all explained samples for each feature.

Kaplan–Meier survival curves were plotted to compare the survival of mortality and several age-related diseases between the low (≤ 65) and high (> 65) SASP Score groups. Differences between groups were evaluated using log-rank tests, with p-values adjusted for multiple testing using the FDR method. The SASP Score cutoff was selected as the 90^th^ percentile of the SASP Score in individuals without a major chronic medical condition at baseline, including cancer (excluding nonmelanoma skin cancer), diabetes (type I diabetes, type II diabetes, and malnutrition-related diabetes), heart failure, myocardial infarction (MI), stroke, chronic obstructive pulmonary disease (COPD), and dementia (ICD-10 codes and field IDs to extract the data in **Table S1**).

For external validation, we calculated Spearman correlation coefficients between the SASP Score and chronological age and medical comorbidity burden, measured by the Cumulative Illness Rating Scale for Geriatrics (CIRS-G) in the MEDEX cohort. Linear mixed-effects models were used to test the effect of exercise on the SASP Score over 18 months, with assessments at baseline, 6 months, and 18 months. Fixed effects included intervention group (exercise vs. non-exercise), time, and their interaction, with a random intercept specified for each participant. P-values associated with the fixed effects were reported. Marginal means were compared across time points within intervention groups, with p-values adjusted for multiple testing using Tukey’s Honestly Significant Difference method.

Statistical significance was defined as p < 0.05. All statistical analyses were performed in R version 4.4.2 using R packages including “survival” (Cox regression models), “survminer” (Kaplan–Meier survival curves), “p.adjust” (multiple testing adjustments), “lme4” (linear mixed effects models), and packages to produce figures such as “forestploter” and “ggplot2”.

## Results

### Baseline characteristics of included UKB-PPP participants (n = 50,997)

Among the included UKB-PPP participants (n=50,997), the mean age was 56.8 years (SD=8.2, range 39 to 70). The majority of participants were of European descent (93.75%) and female (53.98%). Overall, 32.48% of participants held a college or university degree, and the mean level of material deprivation, as measured by the Townsend deprivation index, was lower than the UK population average (−1.18 vs. 0). Nearly half of participants (45.65%) were previous or current smokers, and most of them (95.25%) were previous or current alcohol drinkers. The mean body mass index (BMI) was 27.47 kg/m² (SD=4.80), and the mean waist-to-hip circumference ratio was 0.87 (SD=0.09). The prevalence of hypertension and hypercholesterolemia was 28.19% and 16.02%, respectively. (**Table S2**)

The sample was randomly split into a training set (85%, n=D43,358) and a test set (15%, n=D7,639). Baseline characteristics were comparable between the training and test samples (**Table S3**).

### Correlations between the SASP Score and chronological age, biological aging measures, and aging traits at baseline (test sample, n = 7,639)

Spearman’s correlation coefficients are evaluated between the SASP Score and multiple baseline participant characteristics. SASP Score showed moderate to strong positive correlations with chronological age (rho= 0.70) and other composite biological aging measures, i.e., PhenoAge^29^ (rho = 0.66) and BioAge^30^ (rho = 0.71), proteomic aging clock (PAC)^31^(rho = 0.67) and healthspan proteomic score^32^(HPS) (0–1 scale, lower values indicate worse health; rho = -0.63). In addition, higher SASP Scores showed weak to moderate correlations with higher waist-to-hip circumference ratio (WHR), higher BMI, higher systolic blood pressure, the 49-item frailty index, shorter leukocyte telomere length, lower grip strength, slower walking pace, longer reaction time based on a cognitive function test, and cardiopulmonary fitness (**Fig. S1**).

### SASP Score and common disease risk factors at baseline (test sample, n = 7,639)

At baseline, the median SASP Score was significantly higher in males (vs. females), older adults (≥ 60 years vs. < 60 years), participants with high WHRs (WHR ≥ 0.85 in females and WHR ≥ 0.90 in males), and former or current smokers (vs. non-smokers). Higher median SASP Scores were also associated with frailty (49-item frailty index ≥ 0.21)^33^, hypertension, and hypercholesterolemia (**Fig. 2**).

**Fig. 2.**
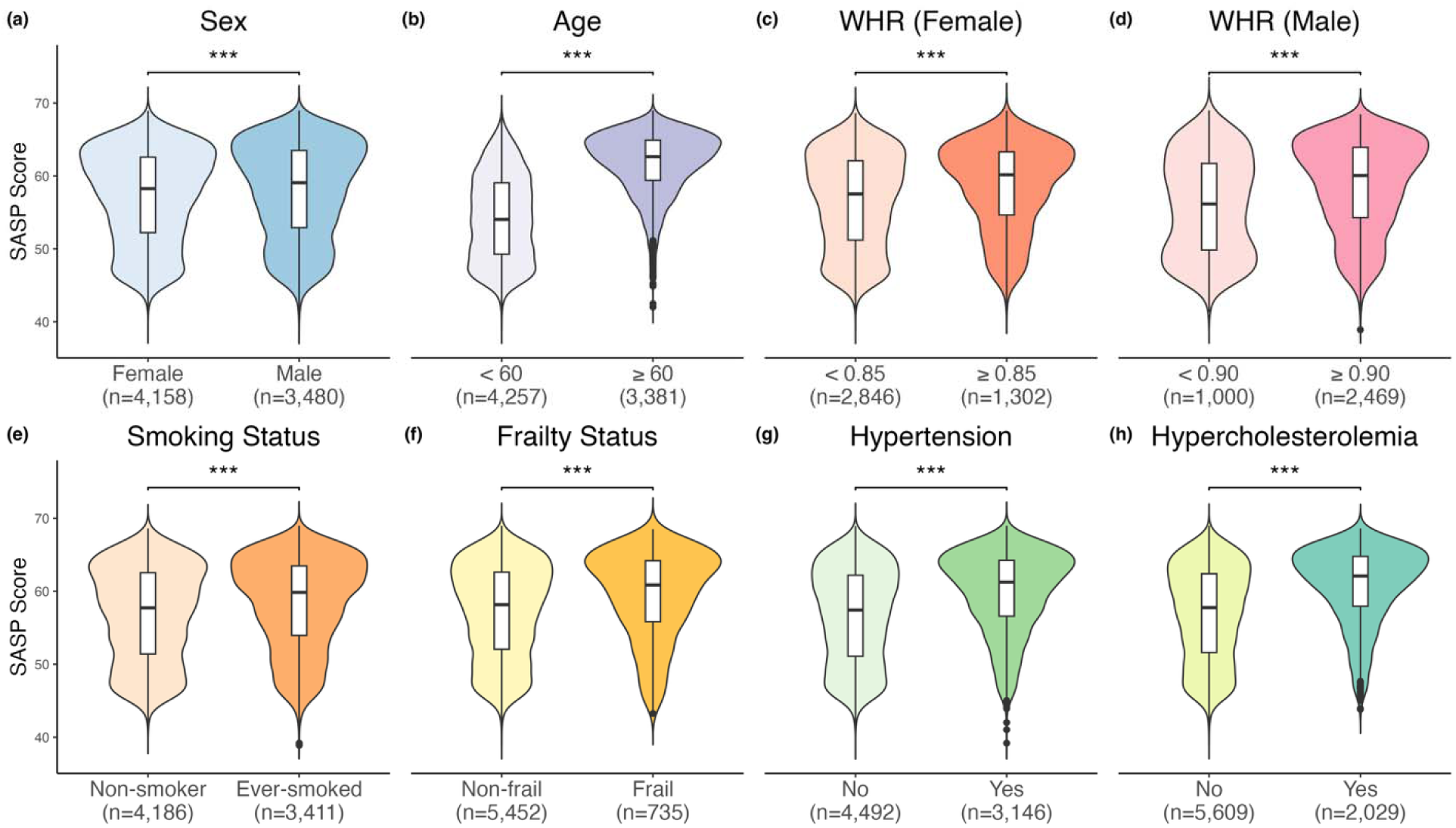
Baseline SASP Score distributions in the UK Biobank test sample (n=7,639) comparing males vs. females (a), older adults vs. younger adults (b), high vs. low waist-to-hip ratio in females (c) and in males (d), previous or current smoker vs. non-smoker (e), frail (49-item frailty index > 0.21) vs. non-frail (f), participants with and without hypertension (g), and participants with and without hypercholesterolemia (h). Wilcoxon rank-sum test p-values adjusted for multiple testing using the Benjamini-Hochberg false discovery rate (FDR) method and are indicated: *pD≤D0.05, **pD≤D0.01, ***pD≤D0.001, NS, not significant.

### SASP Score and the risks of mortality and age-related conditions during follow-up (test sample, n = 7,639)

After controlling for baseline participant characteristics (chronological age, sex, ethnicity, education, Townsend deprivation index, smoking status, alcohol drinker status, systolic blood pressure, and BMI), higher SASP Scores were significantly associated with increased risk of mortality (adjusted hazard ratio (aHR)=1.41, 95% CI 1.25-1.60 per 1 SD) over a mean follow-up of 13.3 years (**Fig. 3**). The SASP Score was also significantly associated with the incidence of multiple age-related diseases, particularly chronic kidney disease, heart failure, lung cancer, and neurological conditions such as delirium, dementia, and Parkinson’s disease (**Fig. 3**). In addition, the SASP Score was robustly predictive of mortality and multiple age-related diseases (**Fig. 4**).

**Fig. 3.**
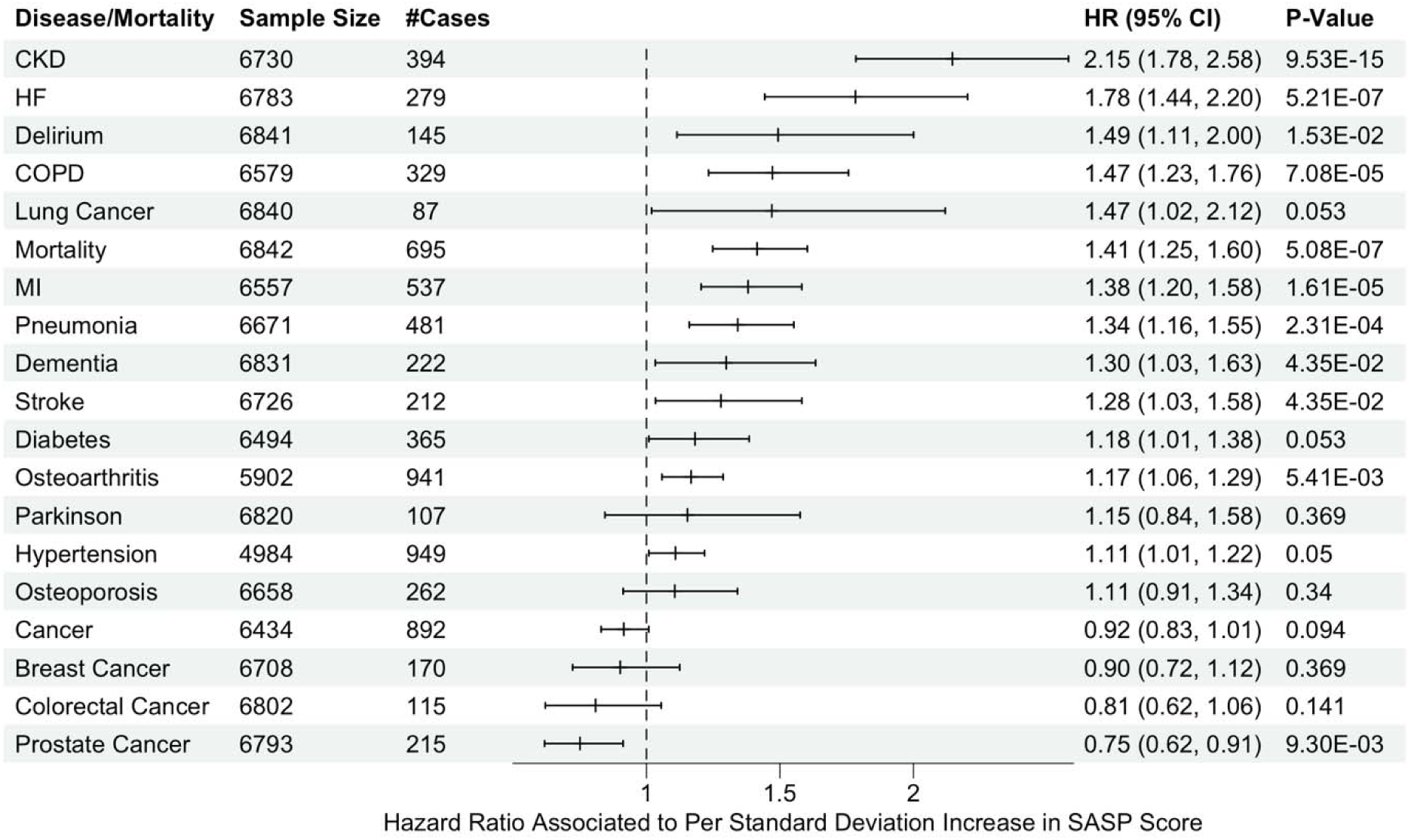
Associations between baseline SASP Score and risk of mortality and age-related diseases in the UK Biobank test sample (n = 7,639). Hazard ratios per 1-SD standard deviation (SD) increase in the SASP Score were estimated using Cox proportional hazards models. Models were adjusted for baseline chronological age, sex, ethnicity, education level, Townsend deprivation index, smoking status, alcohol drinker status, systolic blood pressure, and body mass index (BMI). P-values were adjusted for multiple testing using the Benjamini-Hochberg false discovery rate method.

**Fig. 4.**
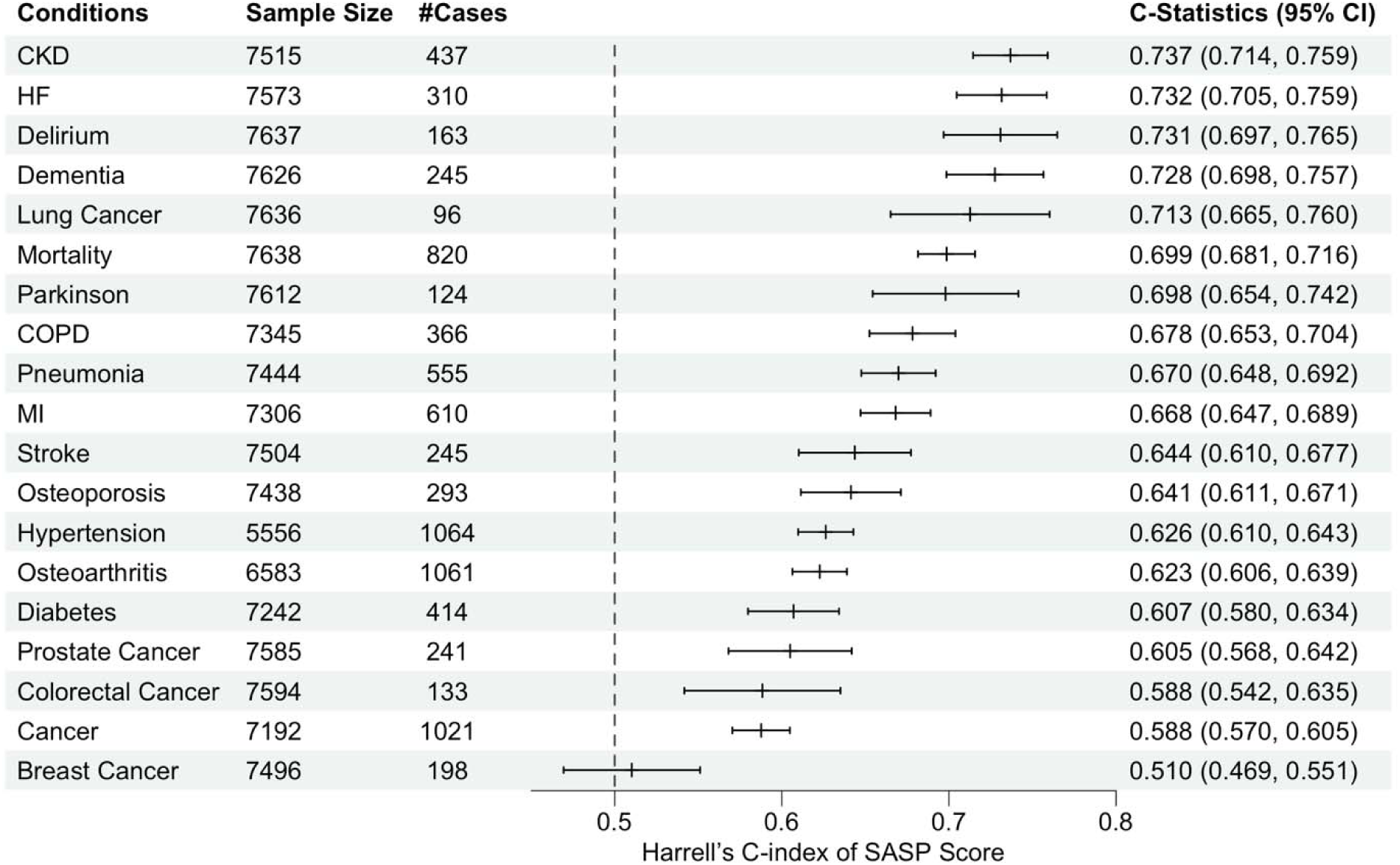
Predictive performance of the SASP Score for time to age-related conditions in the UK Biobank test sample (n = 7,639). Predictive accuracy was evaluated using C-statistics derived from Cox proportional hazards models of the SASP Score for mortality and multiple age-related diseases.

Next, we evaluated the relative contribution of individual proteins to the SASP Score using the Shapley Additive exPlanations (SHAP) analysis^27^. The top 5 contributors to the SASP Score were GDF-15, CXCL-9, PGF, TNFRSF11B, and CCL-20 (**Fig. S3**), highlighting the important roles of mitochondrial function, immune response, and apoptosis control in cellular senescence. We further compared the predictive power of the composite SASP Score with that of its individual components. We found that the SASP Score consistently showed substantially stronger associations with mortality and all age-related diseases (except for diabetes) than any single protein (**Fig. S4-S19**).

### Additional sensitivity analyses (test sample, n = 7,639)

Individuals with high SASP Scores (SASP Score > 65) showed worse survival curves for mortality and several major chronic medical conditions compared to those with low SASP Scores (≤ 65) (**Fig. 5**). The SASP Score cutoff corresponds to the 90^th^ percentile of the SASP Score in the training sample without a major chronic medical condition at baseline, including cancer, diabetes, heart failure, myocardial infarction (MI), stroke, chronic obstructive pulmonary disease (COPD), dementia.

**Fig. 5.**
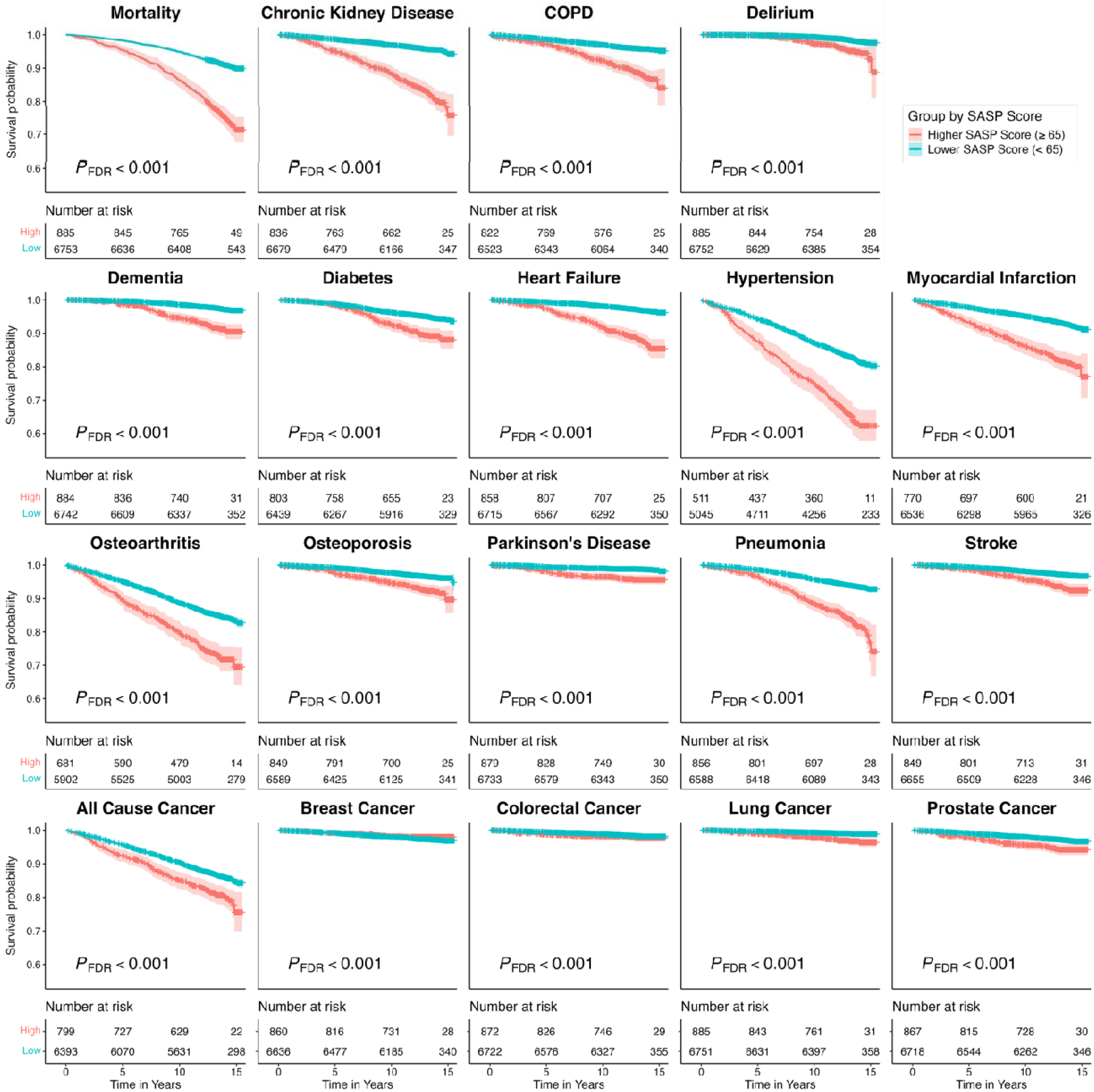
Kaplan–Meier survival curves stratified by SASP Score in the UK Biobank test sample (n = 7,639). Kaplan–Meier curves compare event-free survival for mortality and multiple age-related diseases between participants with high SASP Scores (> 65) and low SASP Scores (≤ 65) at baseline. Time is shown in years from baseline assessment. Shaded areas indicate 95% confidence intervals. P-values were obtained from log-rank tests and adjusted for multiple testing using the Benjamini-Hochberg false discovery rate (FDR) method.

### External validation of the SASP Score model

Using chronological age for fine-tuning guidance, we applied the GAET model to derive the SASP Score in the MEDEX study. The SASP Score showed a modest correlation with age (rho=0.34) and medical comorbidity burden (rho=0.16). The lower correlation in the MEDEX study sample is probably due to its constricted age range (65-84 years at baseline) and that the sample comprises healthier older adults without severe chronic medical conditions (e.g., cardiovascular disease, chronic kidney disease, or cancer).

The linear mixed-effect model showed a significant main effect of time (p<0.001), but no significant effect of intervention (exercise vs. no exercise), or intervention-by-time interaction on the trajectory of SASP Scores (**Fig. 6**). However, the marginal mean comparison of the SASP Score over time within groups revealed an interesting pattern. The SASP Score showed a significant increase from baseline to 6-month follow-up (Tukey adjusted p=0.037) and further from baseline to 18-month follow-up (Tukey adjusted p<0.001) in the non-exercise group, suggesting that the SASP Score increases over time without active exercise intervention. On the other hand, we did not observe a significant increase in the SASP Score from baseline to the 6 or 18-month follow-up in the exercise group. These findings suggest that multi-modal exercise intervention may prevent SASP accumulation in older adults (**Fig. 6**).

**Fig. 6.**
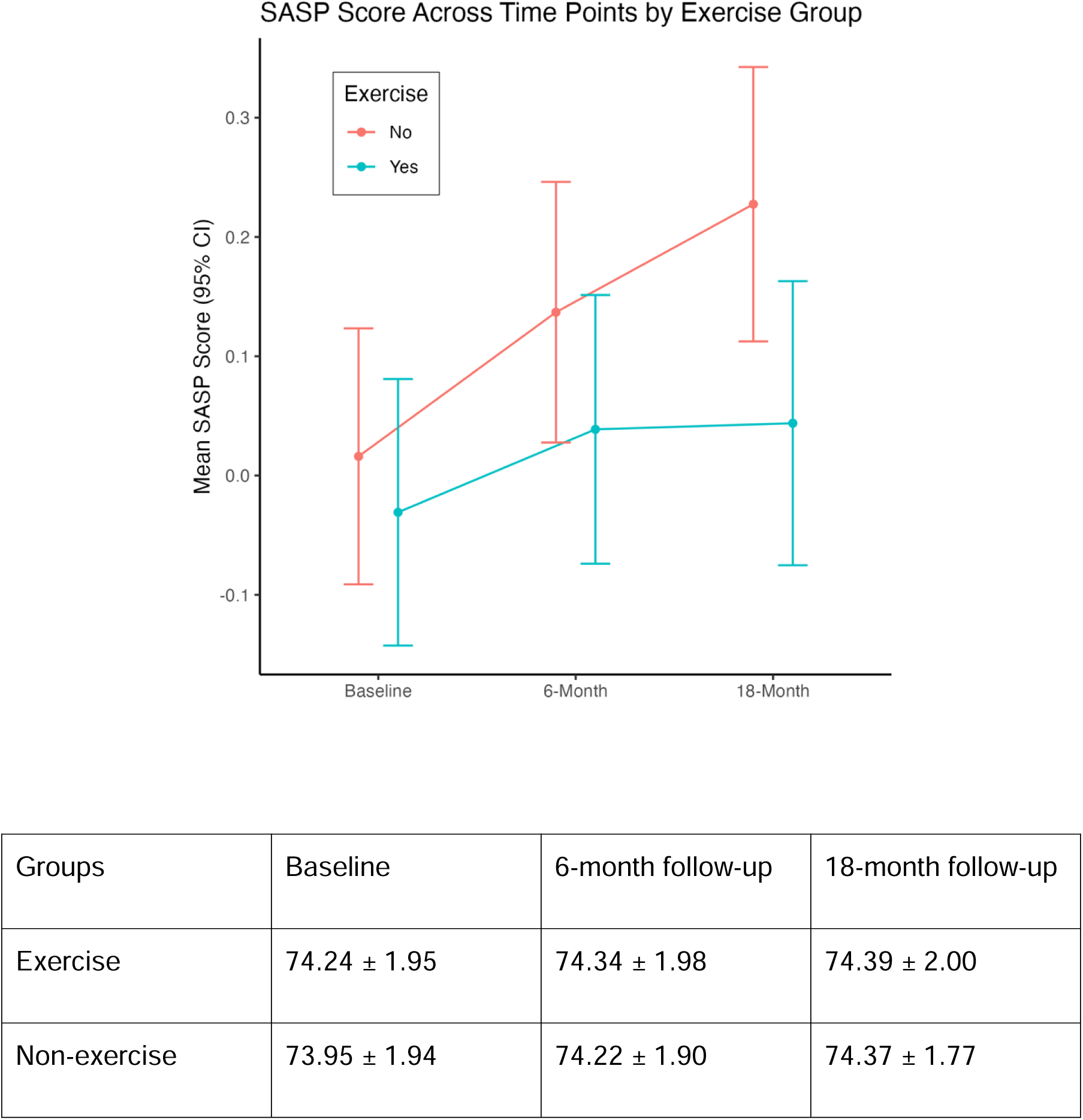
Mean SASP Score trajectories over time by exercise intervention group in the MEDEX cohort (n = 585). Points represent estimated marginal means of the SASP Score at baseline, 6 months, and 18 months for the exercise and non-exercise groups, derived from linear mixed-effects models adjusted for baseline age. Error bars indicate 95% confidence intervals. Within-group changes over time were assessed using marginal mean comparisons with p-values adjusted for multiple testing using Tukey’s Honestly Significant Difference method.

## Discussion

In the present study, we developed a composite biomarker based on proteins commonly associated with the secretome of senescent cells, the SASP Score, using a deep learning framework, the guided autoencoder with transformer (GAET). The SASP Score showed strong predictive power and prognostic accuracy to identify individuals at higher risk of developing different medical chronic conditions and death. We derived the SASP Score in an independent cohort and showed that it significantly increases over time, but exercise can stabilize its levels over 18 months. Our results suggest that the SASP Score can be used as a prognostic biomarker in observational studies and in clinical trials to evaluate the effect of interventions targeting cellular senescence (i.e., senolytics) or SASP (e.g., senomorphics).

The search for biomarkers that can serve as prognostic tools and evaluate the biological effects of different interventions is an area of intense investigation^34^. Several proteomic-based models have been developed recently, mainly to assess systemic or organ-specific biological aging acceleration^31,35,36^ or healthspan^32^. Despite their robustness, the widespread implementation of these models may be limited, as their use across studies depends on data from the same proteomic platform on which they were developed. The SASP Score has overcome this limitation by applying a novel semi-supervised deep learning model, the guided autoencoder with transformer. The GAET, by combining the powerful pattern modeling of transformers with the data compression of autoencoders, can reduce high-dimensional data (i.e., the individual SASP proteins) into a concise, feature-rich representation (i.e., the SASP Score) that can be used for downstream analyses. Moreover, the fine-tuning process implemented by the GAET model can also address the age bias commonly observed across different biological aging models^37^.

In this study, we showed the SASP Score, developed using proteomic data from a proximity extension assay in the UK Biobank, can be derived using proteomic data from a multiplex immunoassay in an independent validation study. This translation has major implications as the SASP Score can be calculated using data generated across different proteomic platforms, enabling the evaluation of SASP biomarkers in secondary analyses of multiple studies and potentially reducing costs in future observational studies and clinical trials. Also, it is important to note that the SASP Score had significantly greater predictive power for adverse health outcomes (e.g., mortality) than individual SASP proteins. A possible explanation is that the GAET model, by incorporating non-linear interaction among the SASP proteins, can better capture the complexity of the cellular senescence secretome. Thus, it provides a more comprehensive measurement of the SASP and its relationship to adverse health outcomes, or how it changes after interventions.

Exercise and physical activity has long been established as a health-promoting intervention and widely recommended for primary and secondary prevention of many chronic medical conditions^38,39^. In the external validation cohort, we found that the multimodal exercise intervention did not reduce the SASP score but appeared to prevent the expected age-related increase in the SASP Score over 18 months (**Fig. 6**). Our results are consistent with a recent clinical trial that included 1,377 older adult participants, showing that a physical activity intervention did not significantly reduce plasma levels of individual SASP biomarkers compared with healthy aging education over 24 months, despite the association of higher physical activity levels and lower levels of several SASP proteins^40^. In contrast, a smaller study, including 34 older adults, showed that endurance and strength exercise decreased SASP levels, calculated based on a partial least squares discriminant analysis, over a short intervention period (12 weeks)^41^. Interestingly, a similar pattern of changes in aging biomarkers (clinical chemistry aging clocks, epigenetic aging clocks, and individual SASP proteins) has also been observed in a long-term study of caloric restriction in older adults^42–44^. Altogether, these results suggest that physical activity may modulate and stabilize the SASP levels over time, indicating that it can prevent the accumulation of cellular senescence burden in older adults.

It is important to note that the SASP Score should be viewed as a composite measure developed to evaluate and track changes in the SASP, serving as a proxy for cellular senescence burden at the systemic level. In other words, it is a measure of a specific hallmark of biological aging (i.e., cellular senescence)^1^ rather than a biological aging clock, such as the proteomic aging clock (PAC)^31^, ProtAge^35^, organ-specific proteomic clocks^45^, or the healthspan proteomic score (HPS)^32^. Therefore, the SASP Score can be used in conjunction with other biological aging clocks or measures and provide complementary information about the state of biological aging processes in an individual. Moreover, the SASP Score can be useful in studies evaluating the impact of cellular senescence burden on distinct health outcomes, as well as the effects of non-pharmacological (e.g., lifestyle modifications) and pharmacological (e.g., senolytics or senomorphic drugs) interventions on cellular senescence burden and secretome.

A potential limitation of our study is the choice of the SASP biomarkers. There is no consensus on which SASP markers are universal^6^, and we selected SASP proteins that exhibit similar behaviors across different cell types and senescence stimuli, based on multiple databases and manual curation. Several other studies investigated smaller panels of SASP proteins^8,9,46^; however, they did not analyze them as composite biomarker indices, but instead focused on individual markers and their association with adverse outcomes. One major advantage of analyzing individual proteins and their relation to health outcomes is the ease of interpretation.

However, such an approach carries a higher risk of spurious statistical associations due to inflated type I statistical error and does not capture the complex interactions between proteins, or the heterogeneity of cellular senescence in living organisms. Our SASP Score, on the other hand, provides major advances by integrating multiple proteins relevant to cellular senescence into a composite score that captures complex, non-linear relationships among SASP proteins. Thus our approach is superior to analyzing individual proteins since we can reduce the risk of spurious associations by limiting multiple testing, while capturing the complexity of cellular senescence phenomena. Finally, the GAET model enables the calculation of the SASP Score using data from distinct proteomics platforms. This represents a major advantage in environments with limited financial or lab resources, or in secondary data analysis, where proteomic assay selection is constrained.

In conclusion, we developed a deep learning-based model of the senescence-associated secretory phenotype, i.e., the SASP Score, and showed that it has greater validity and performance compared to a single marker. This approach is also superior to conventional methods in capturing complex relationships within biological process relevant to aging. Future studies should focus on validating our model in different cohorts and evaluating the impact of geroscience-guided interventions on SASP Score trajectories.

## Data Availability

Data access to the UK Biobank is granted upon application.
To request consideration for access to MEDEX study data, please contact Michelle Voegtle, Study Coordinator, via.

## Acknowledgements

This research has been conducted using the UK Biobank Resource under application number 92647 (PI: Richard H. Fortinsky). The study uses data provided by patients and collected by the UK National Health Service (NHS) as part of their care and support.

## Authors’ contributions

CLK and BSD designed the study. SZ and TH developed the SASP Score. SZ conducted the statistical analyses with guidance from CLK and BSD. SZ, CLK, and BSD drafted the manuscript, and all co-authors reviewed and approved the final version.

## Data, Materials, and Software Availability

Data access to the UK Biobank is granted upon application, The Python code can be obtained from the GitHub repositories at https://github.com/shangshuz/sasp. This GitHub repository contains the code for model training, the pre-trained model weights, and the environment specification. All statistical analyses were performed using R version 4.4.2.

## Funding Information

SZ, CLK, BSD, RHF, and GAK: NIA P30AG067988.

EJL, JW, BSD: NIA R01AG072694

## Supplementary Figures

**Fig. S1.**
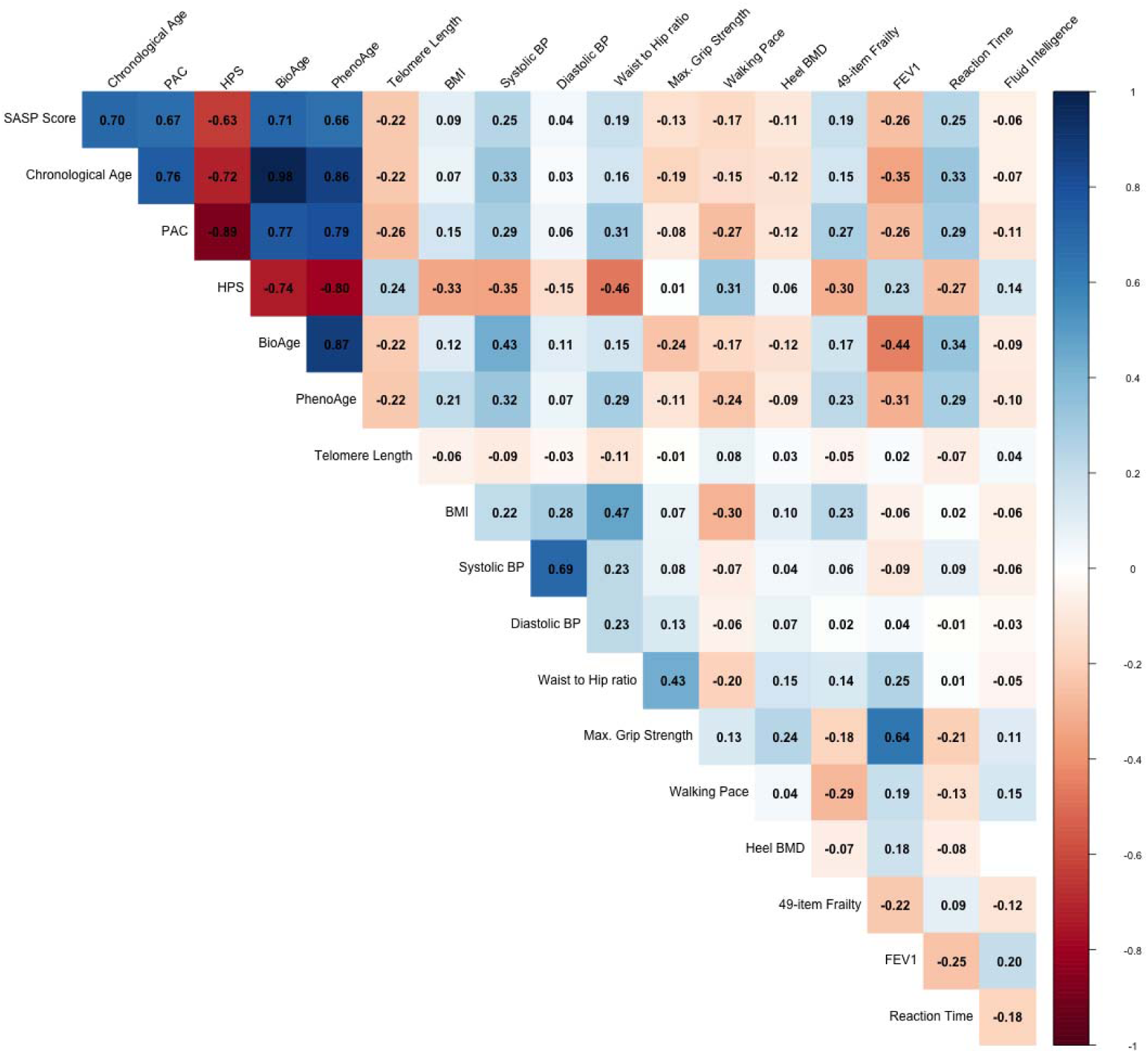
[UKB test samples (n=7,639)] SASP score Spearman correlations with chronological age, other biological age measures, physiological, cognitive, and frailty measures at baseline.

**Fig. S2.**
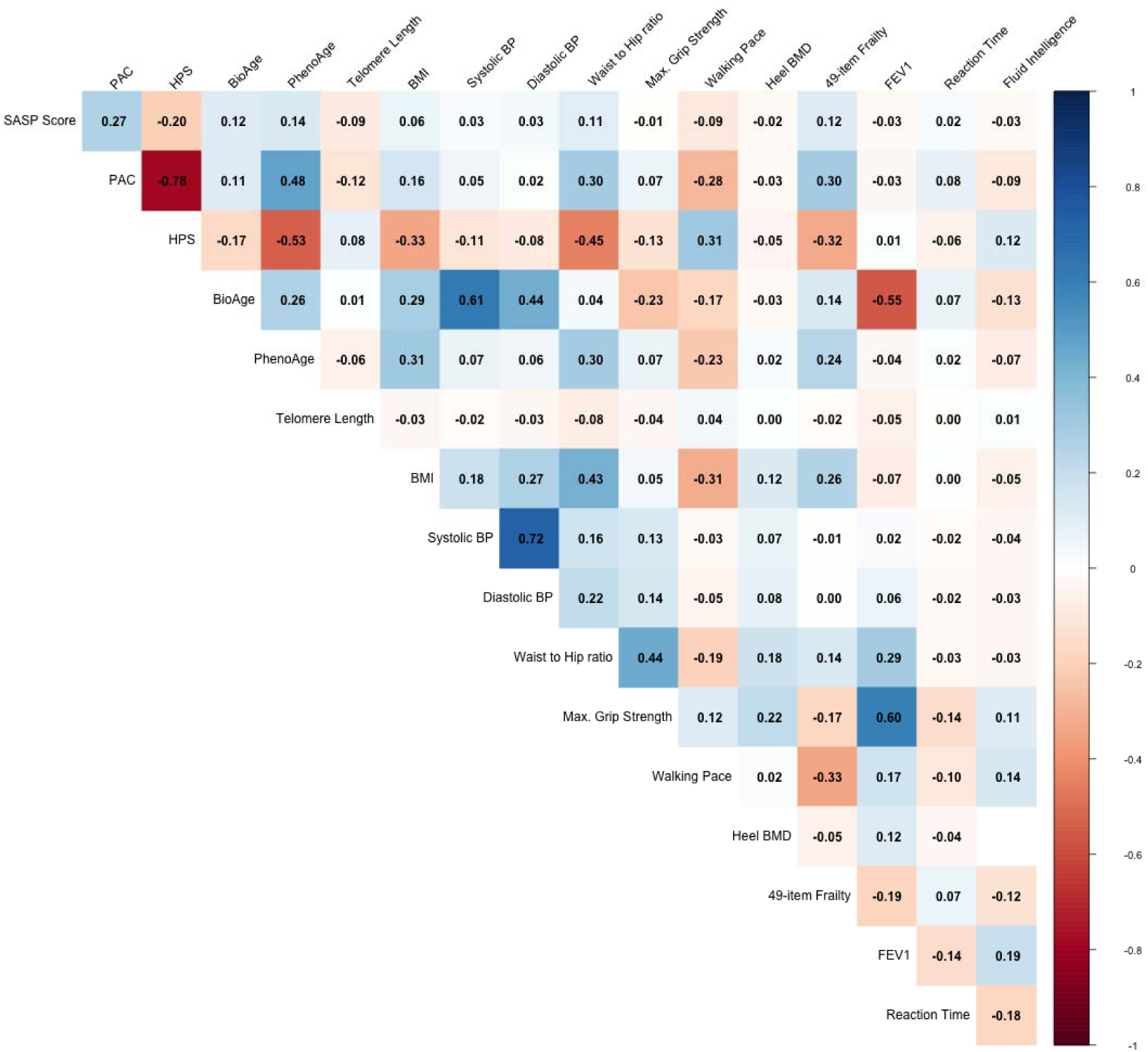
[UKB test samples (n=7,639)] Partial Spearman correlations between SASP score and biological age measures, physiological, cognitive, and frailty measures at baseline, controlling for chronological age.

**Fig. S3.**
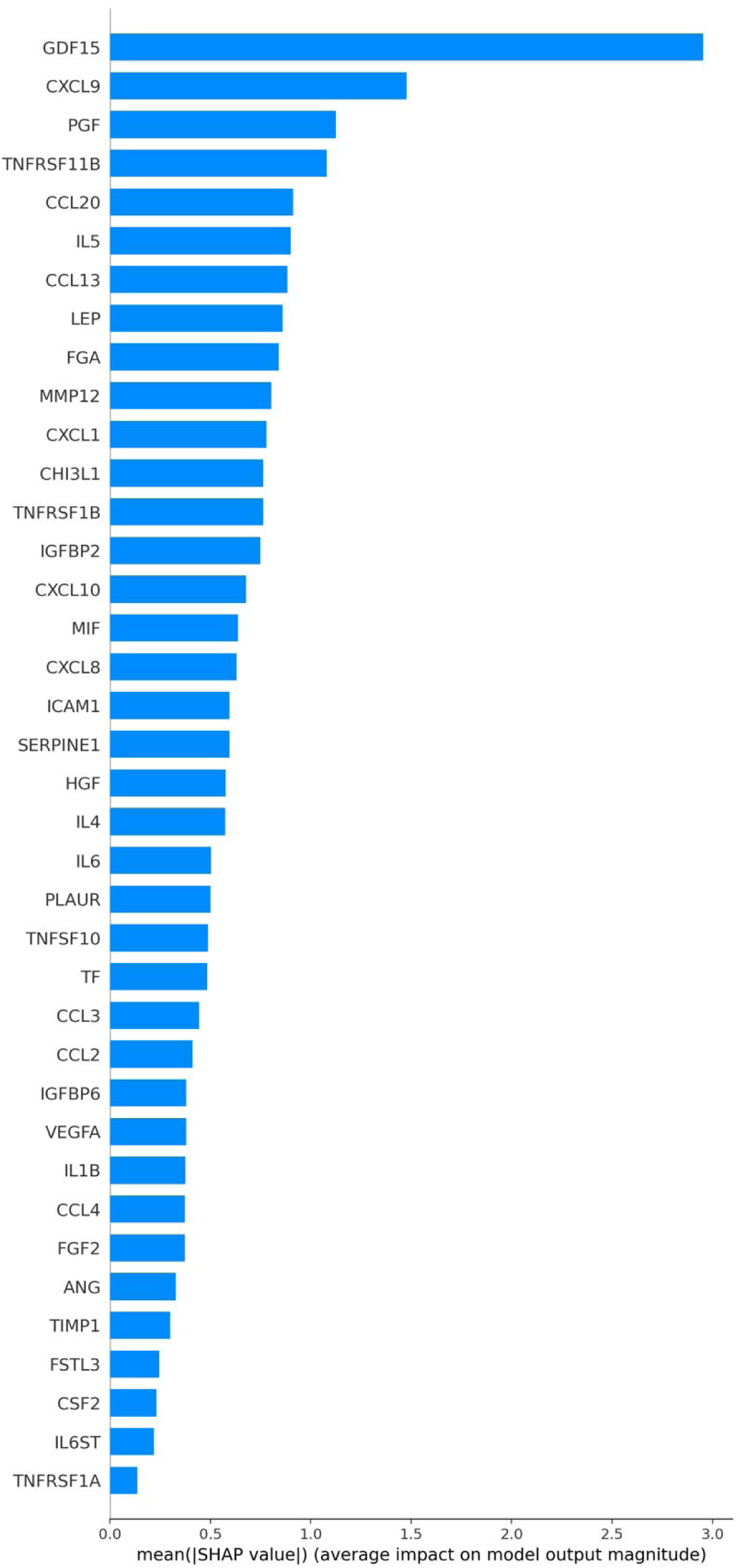
[UKB test samples (n=7,639)] Rank of relative contribution of individual proteins to the SASP Score. Contributions evaluated by mean absolute SHAP value across all explained samples for each protein.

**Fig. S4.**
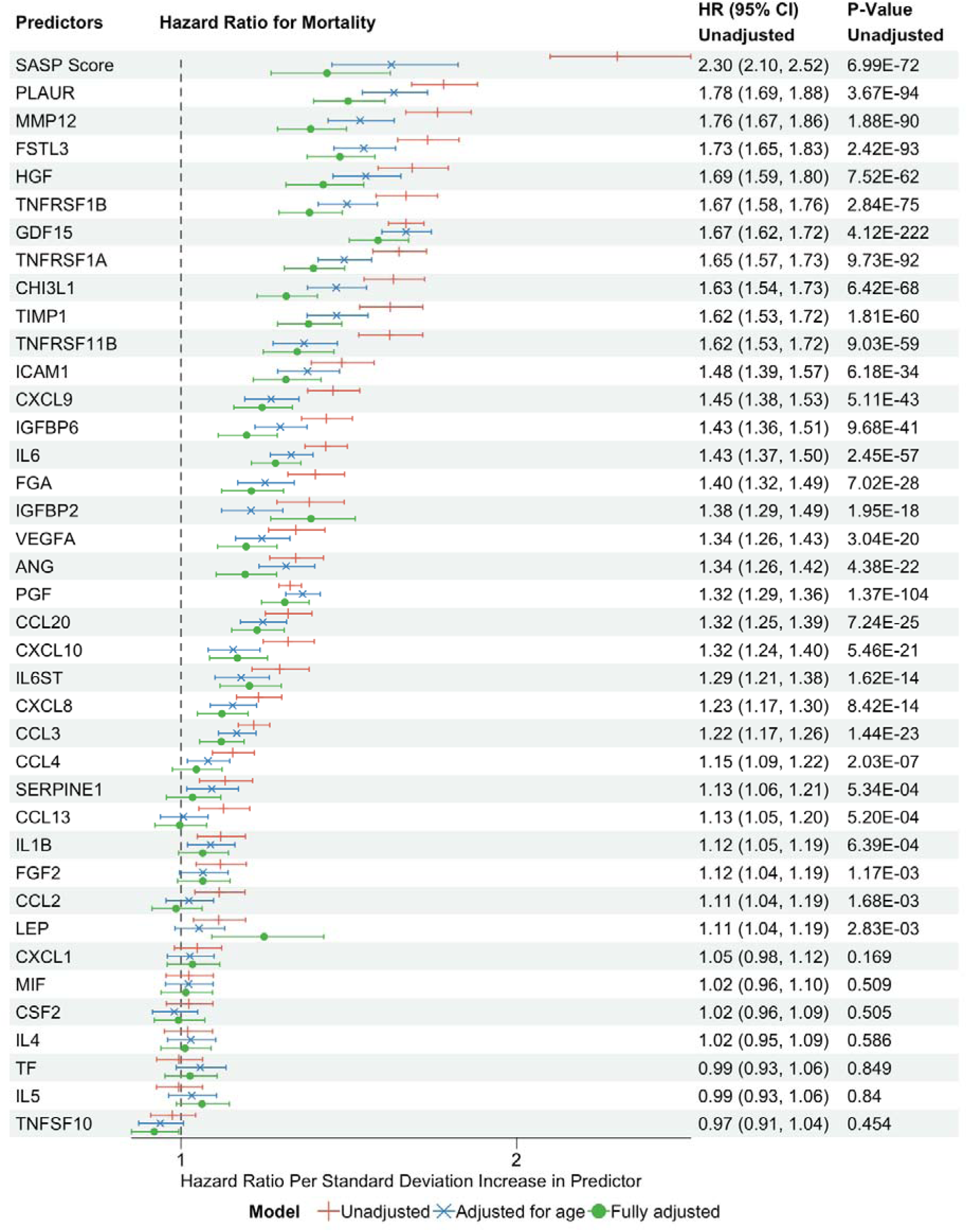
[UKB test samples (n=7,639)] Hazard ratio of mortality comparing SASP score to individual SASP proteins at baseline. Adjusted covariates: age, sex, ethnicity, BMI, education, Townsend, smoking status, systolic blood pressure.

**Fig. S5.**
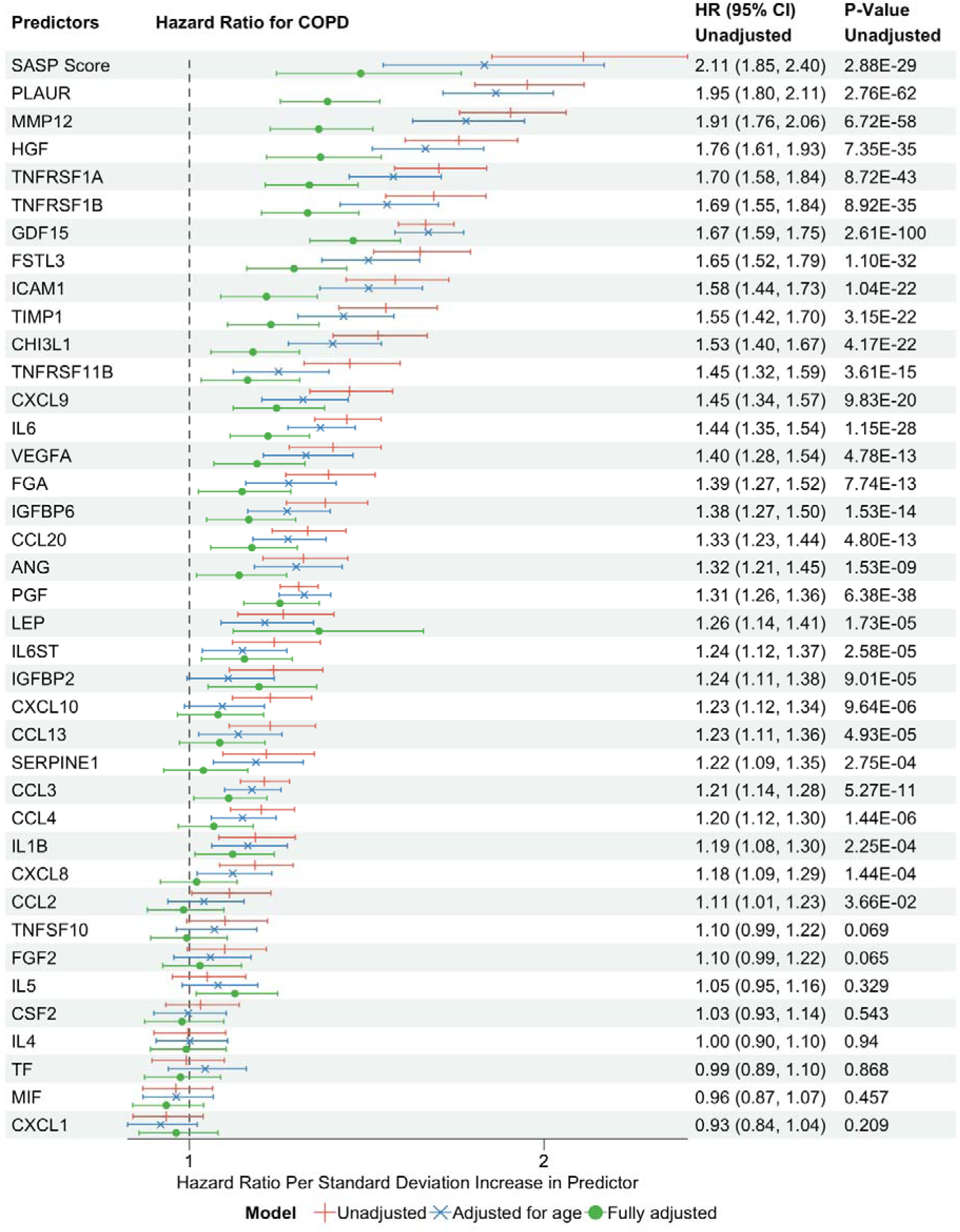
[UKB test samples (n=7,639)] Hazard ratio of COPD comparing SASP score to individual SASP proteins at baseline. Adjusted covariates: age, sex, ethnicity, BMI, education, Townsend, smoking status, systolic blood pressure.

**Fig. S6.**
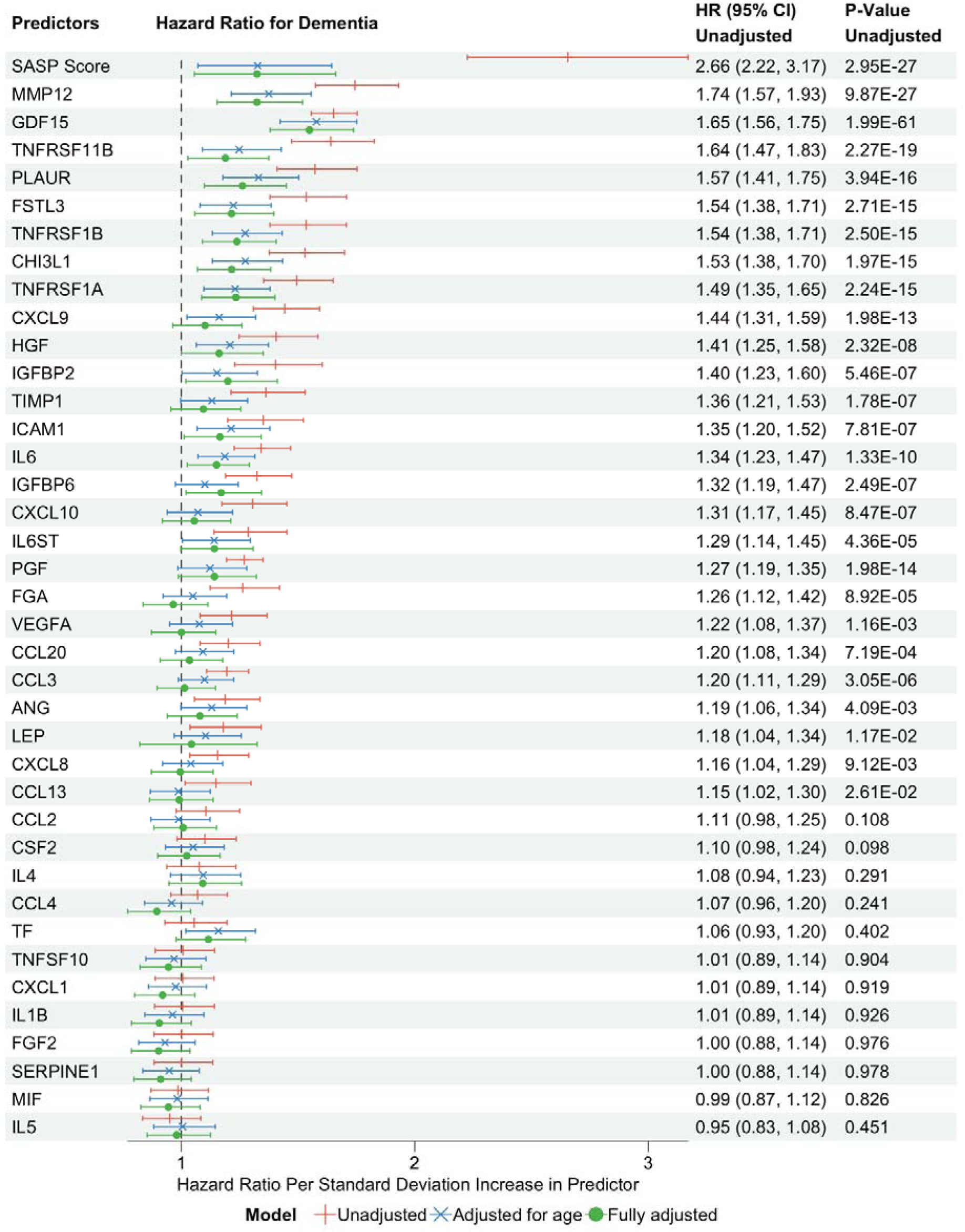
[UKB test samples (n=7,639)] Hazard ratio of dementia comparing SASP score to individual SASP proteins at baseline. Adjusted covariates: age, sex, ethnicity, BMI, education, Townsend, smoking status, systolic blood pressure.

**Fig. S7.**
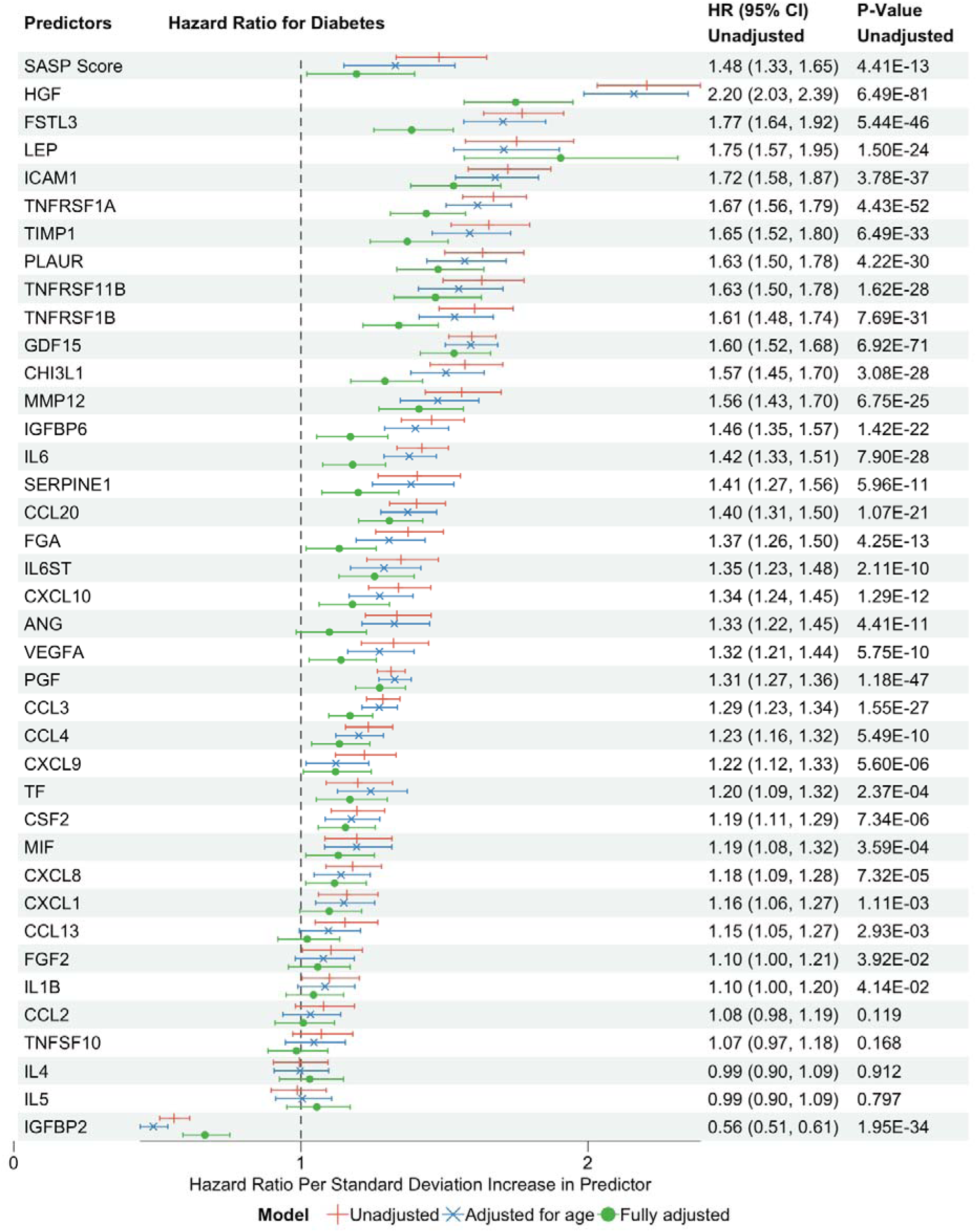
[UKB test samples (n=7,639)] Hazard ratio of diabetes comparing SASP score to individual SASP proteins at baseline. Adjusted covariates: age, sex, ethnicity, BMI, education, Townsend, smoking status, systolic blood pressure.

**Fig. S8.**
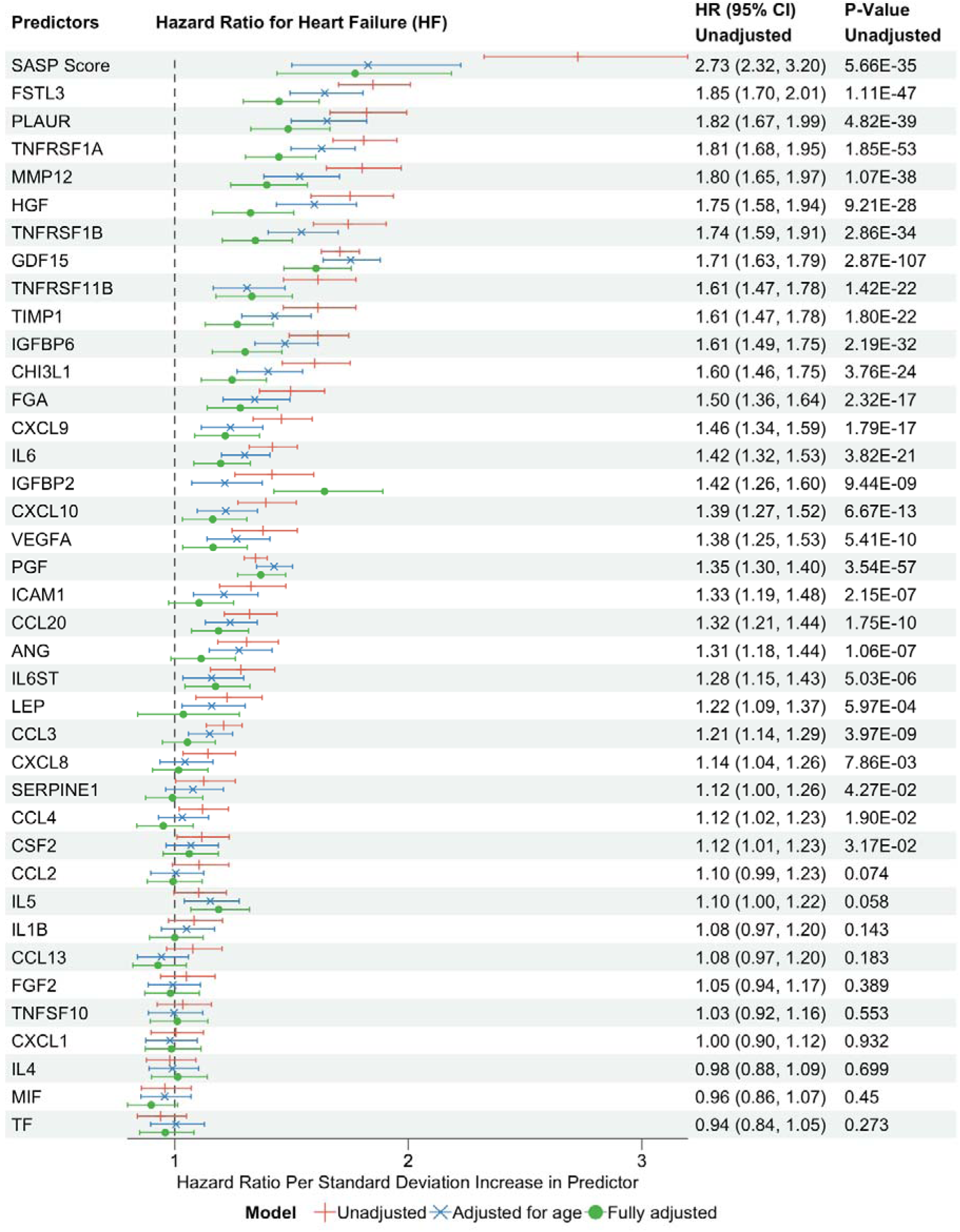
[UKB test samples (n=7,639)] Hazard ratio of heart failure comparing SASP score to individual SASP proteins at baseline. Adjusted covariates: age, sex, ethnicity, BMI, education, Townsend, smoking status, systolic blood pressure.

**Fig. S9.**
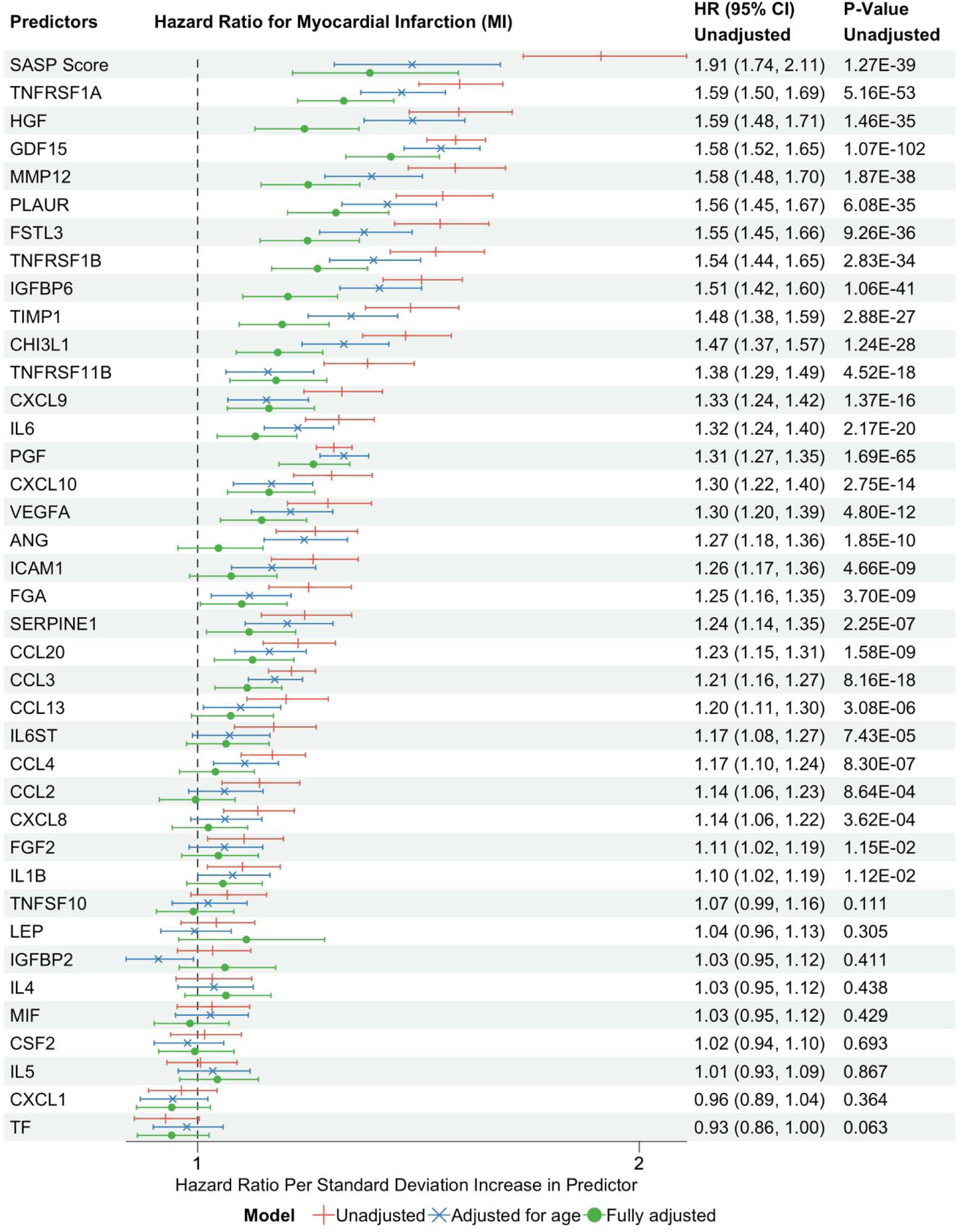
[UKB test samples (n=7,639)] Hazard ratio of MI comparing SASP score to individual SASP proteins at baseline. Adjusted covariates: age, sex, ethnicity, BMI, education, Townsend, smoking status, systolic blood pressure.

**Fig. S10.**
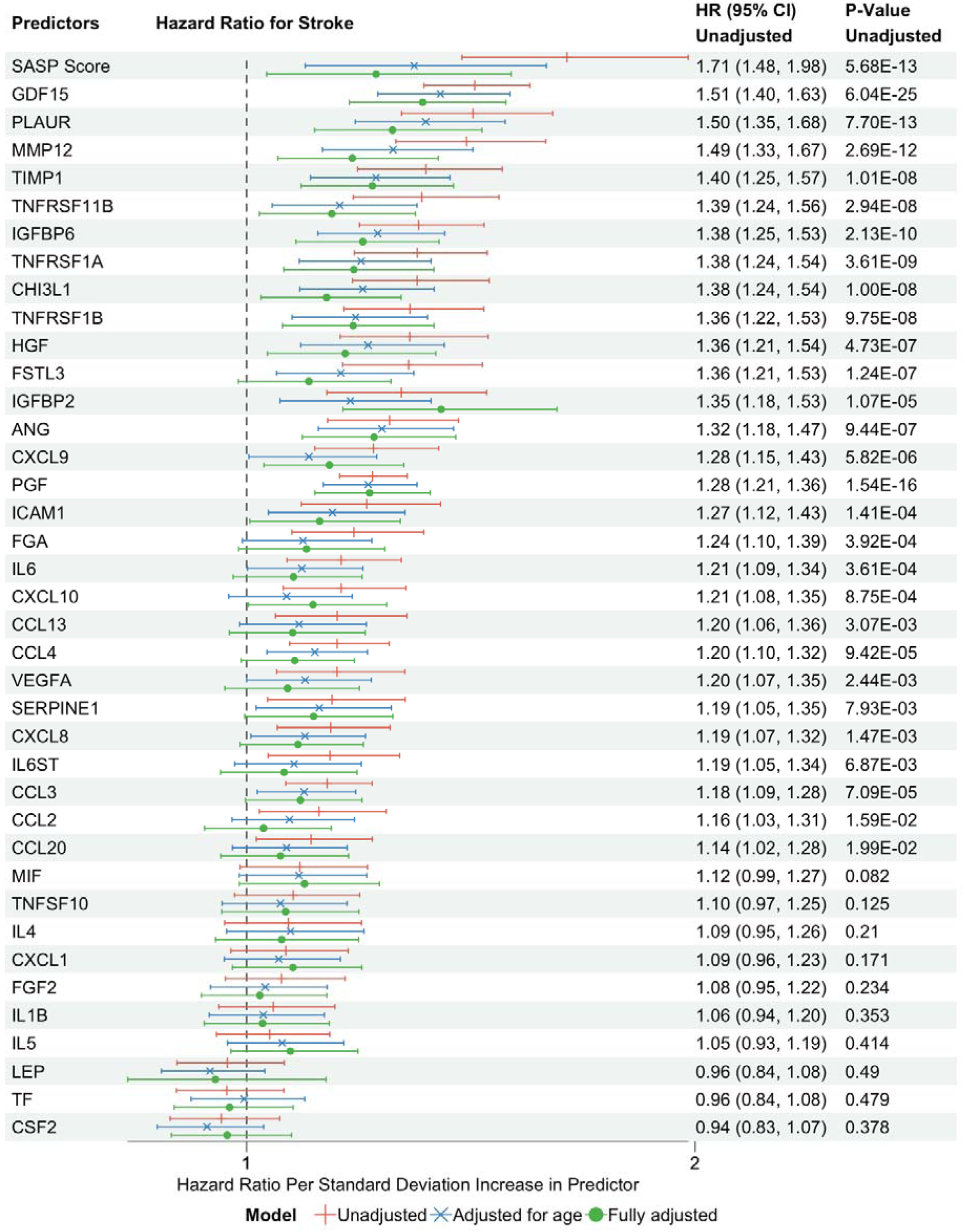
[UKB test samples (n=7,639)] Hazard ratio of stroke comparing SASP score to individual SASP proteins at baseline. Adjusted covariates: age, sex, ethnicity, BMI, education, Townsend, smoking status, systolic blood pressure.

**Fig. S11.**
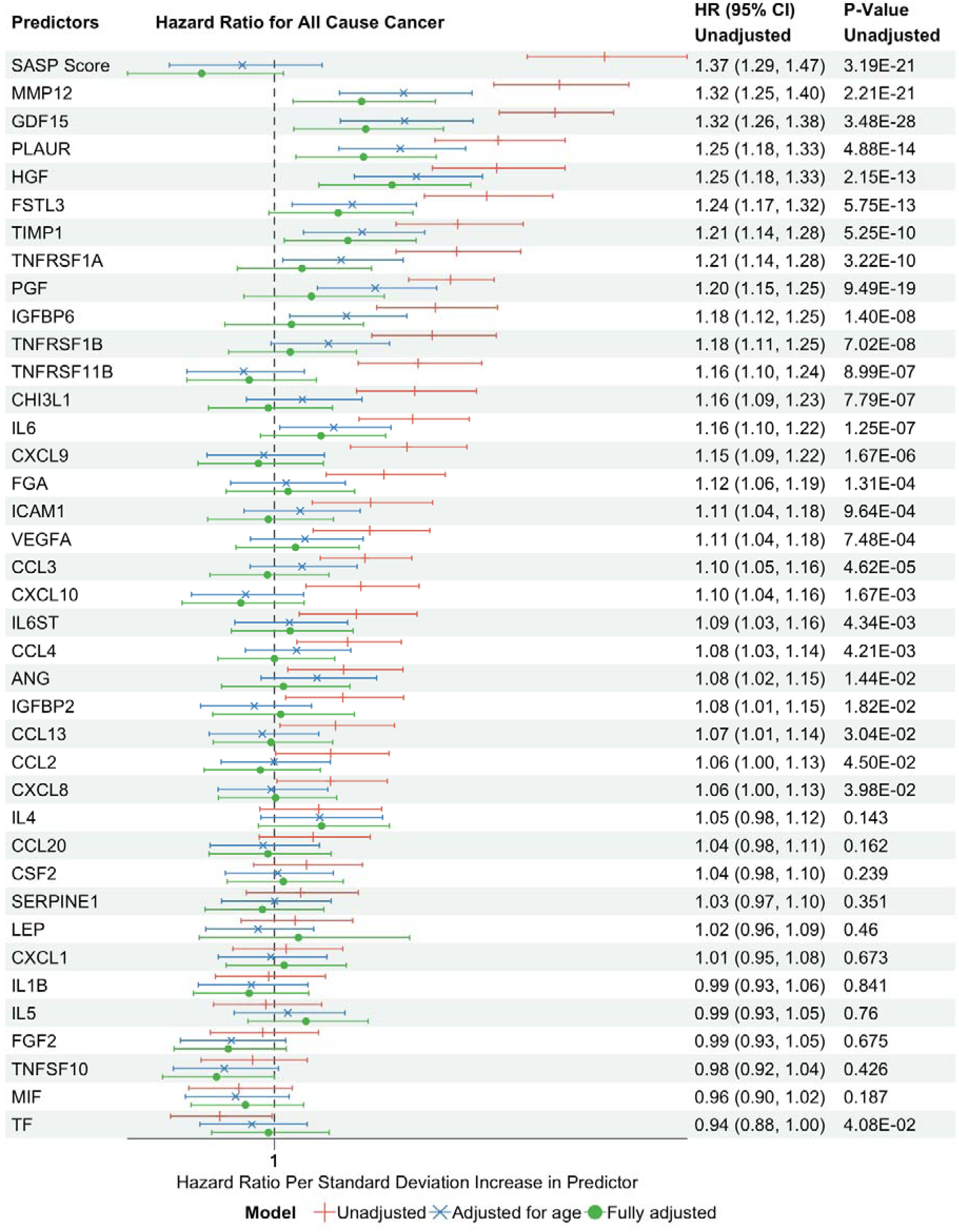
[UKB test samples (n=7,639)] Hazard ratio of all cause cancer comparing SASP score to individual SASP proteins at baseline. Adjusted covariates: age, sex, ethnicity, BMI, education, Townsend, smoking status, systolic blood pressure.

**Fig. S12.**
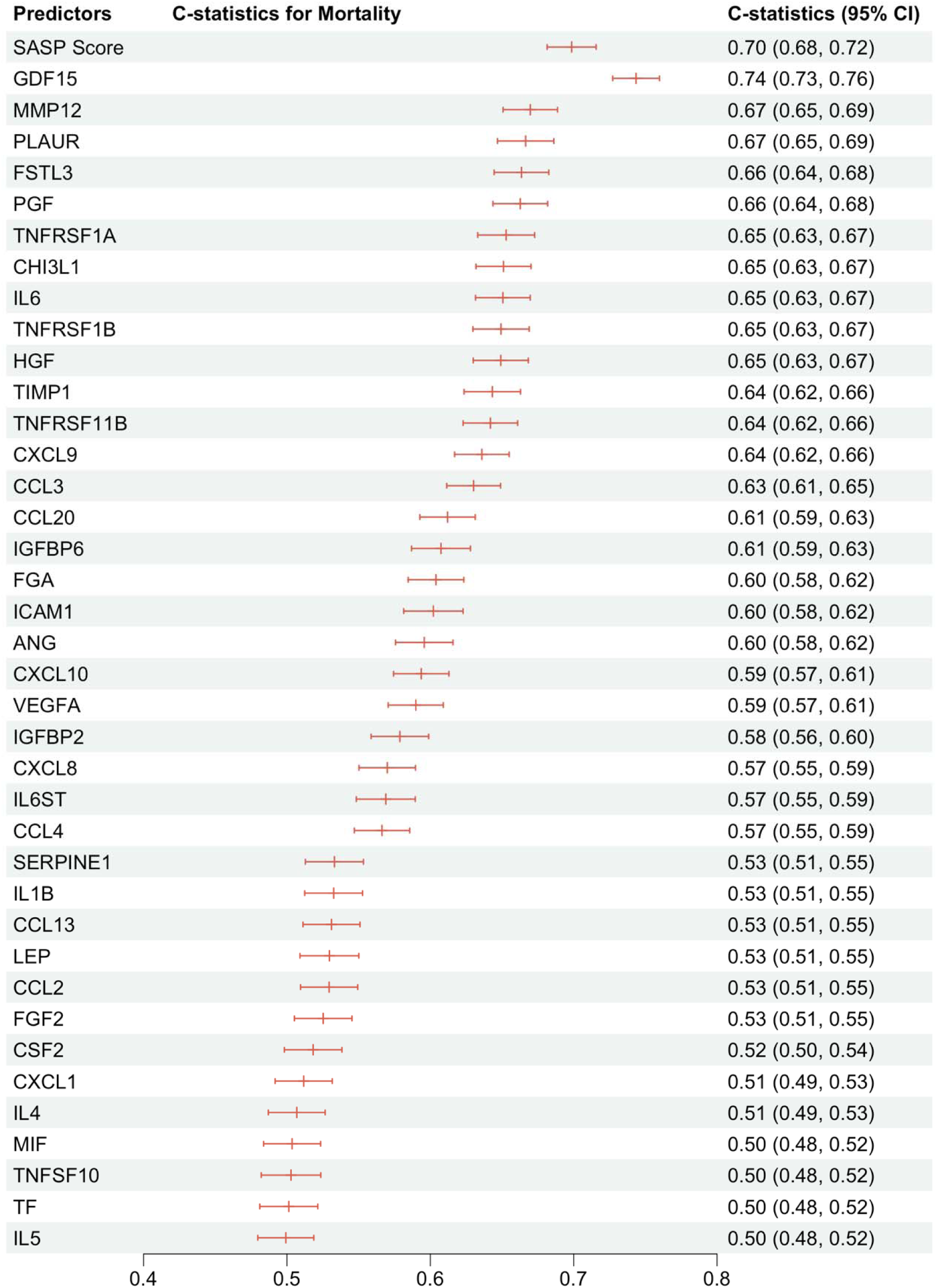
[UKB test samples (n=7,639)] C-statistics for mortality predicted by baseline SASP score and individual proteins.

**Fig. S13.**
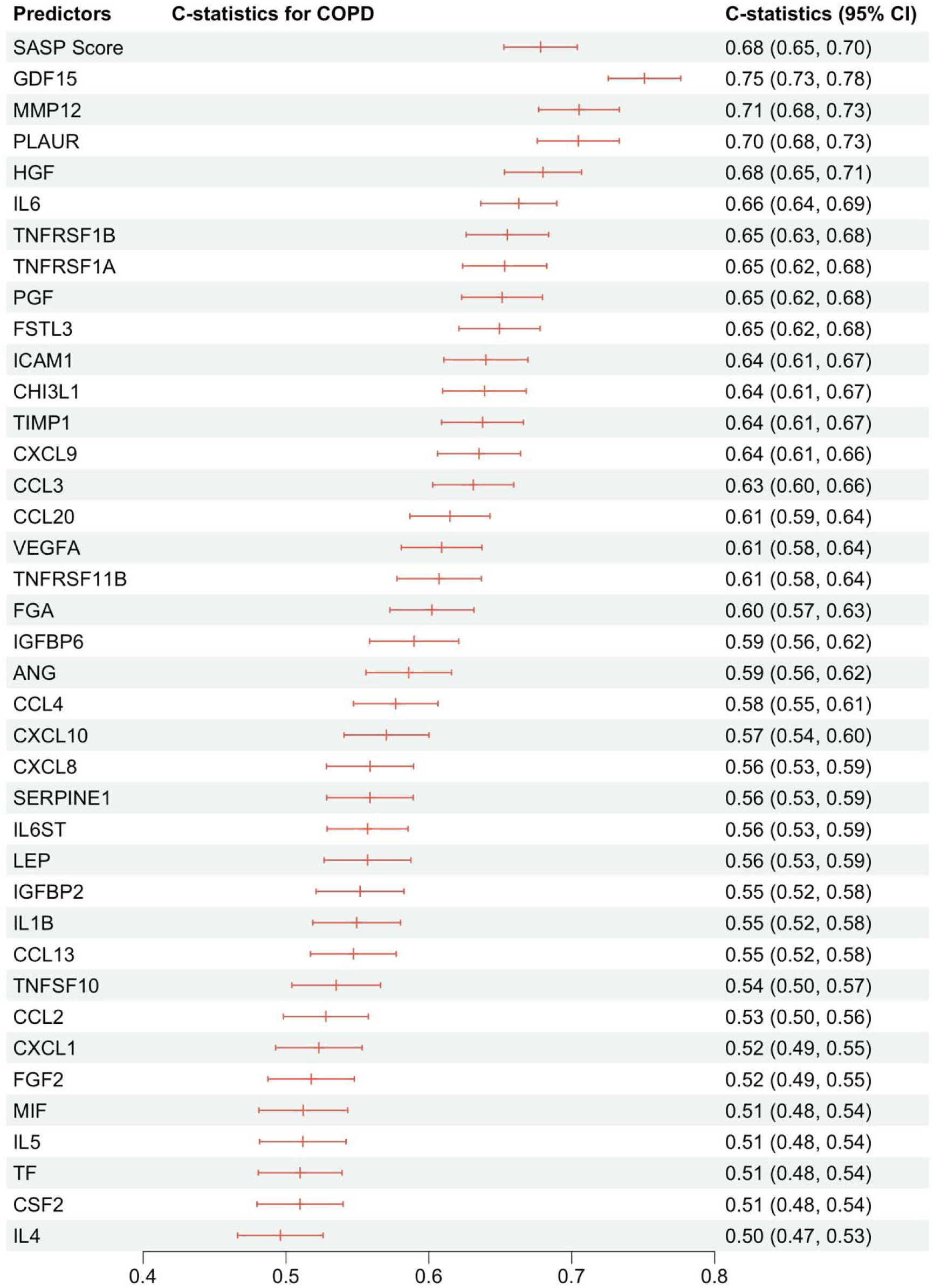
[UKB test samples (n=7,639)] C-statistics for incident COPD predicted by baseline SASP score and individual proteins.

**Fig. S14.**
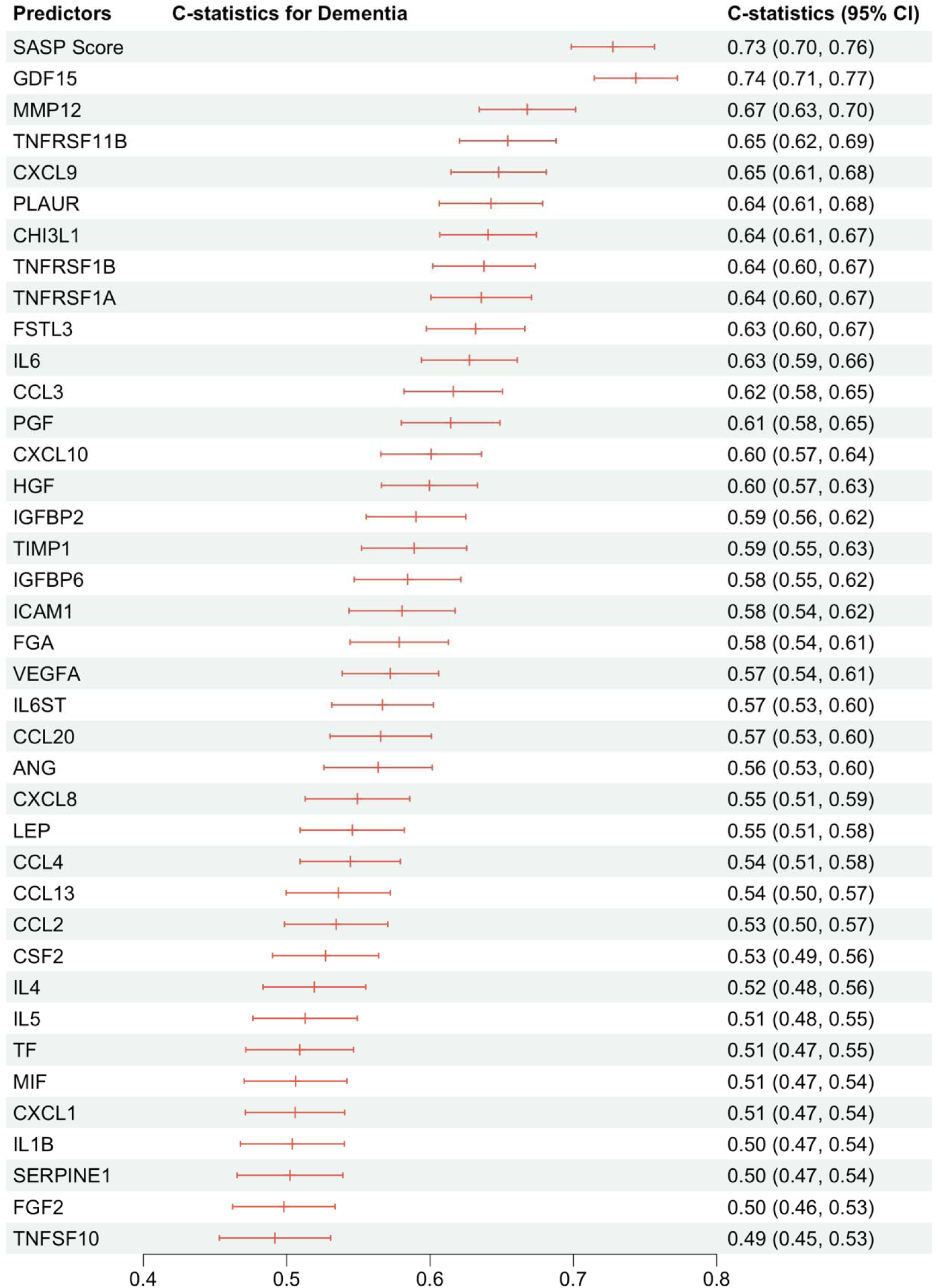
[UKB test samples (n=7,639)] C-statistics for incident dementia predicted by baseline SASP score and individual proteins.

**Fig. S15.**
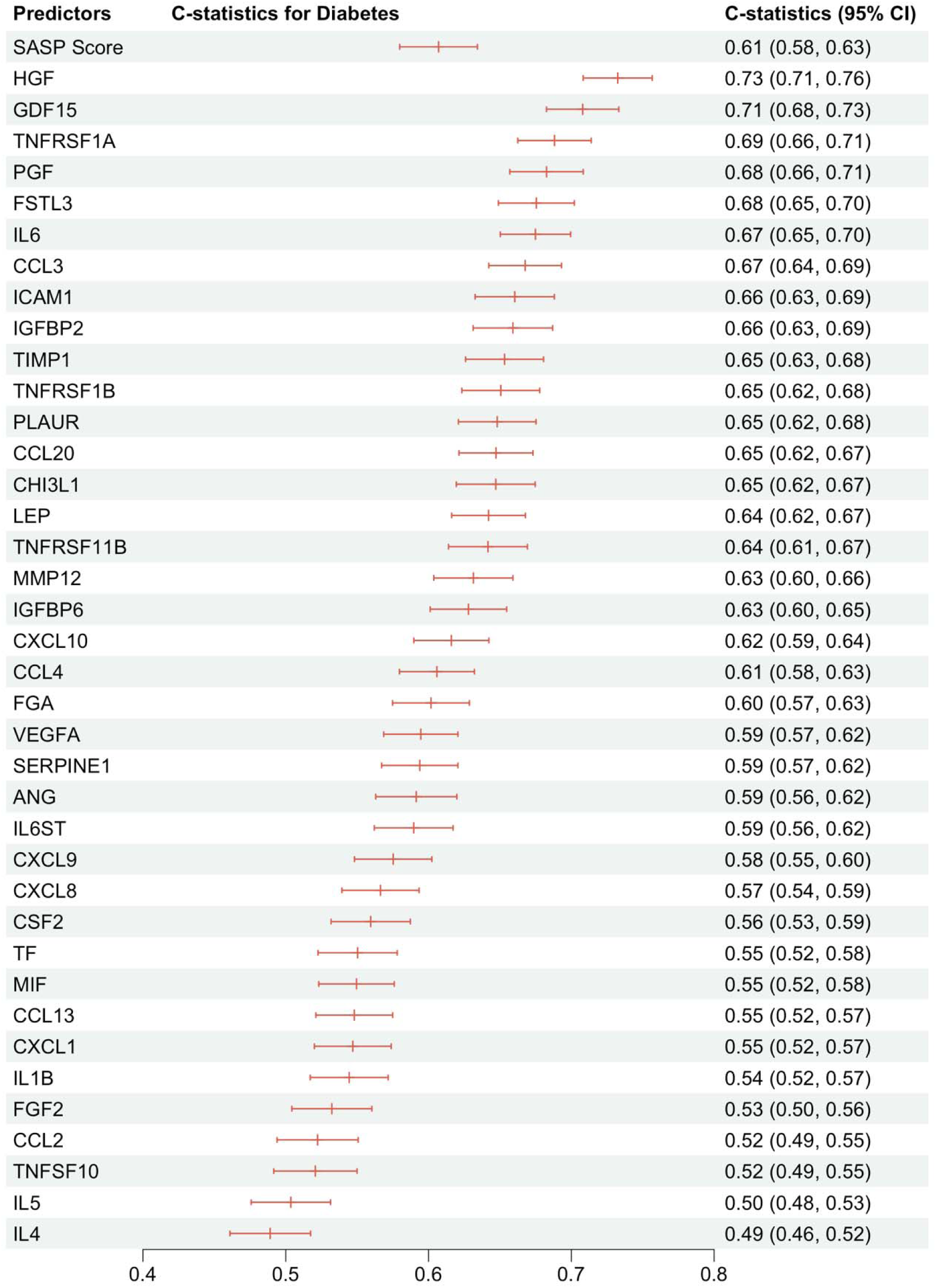
[UKB test samples (n=7,639)] C-statistics for incident diabetes predicted by baseline SASP score and individual proteins.

**Fig. S16.**
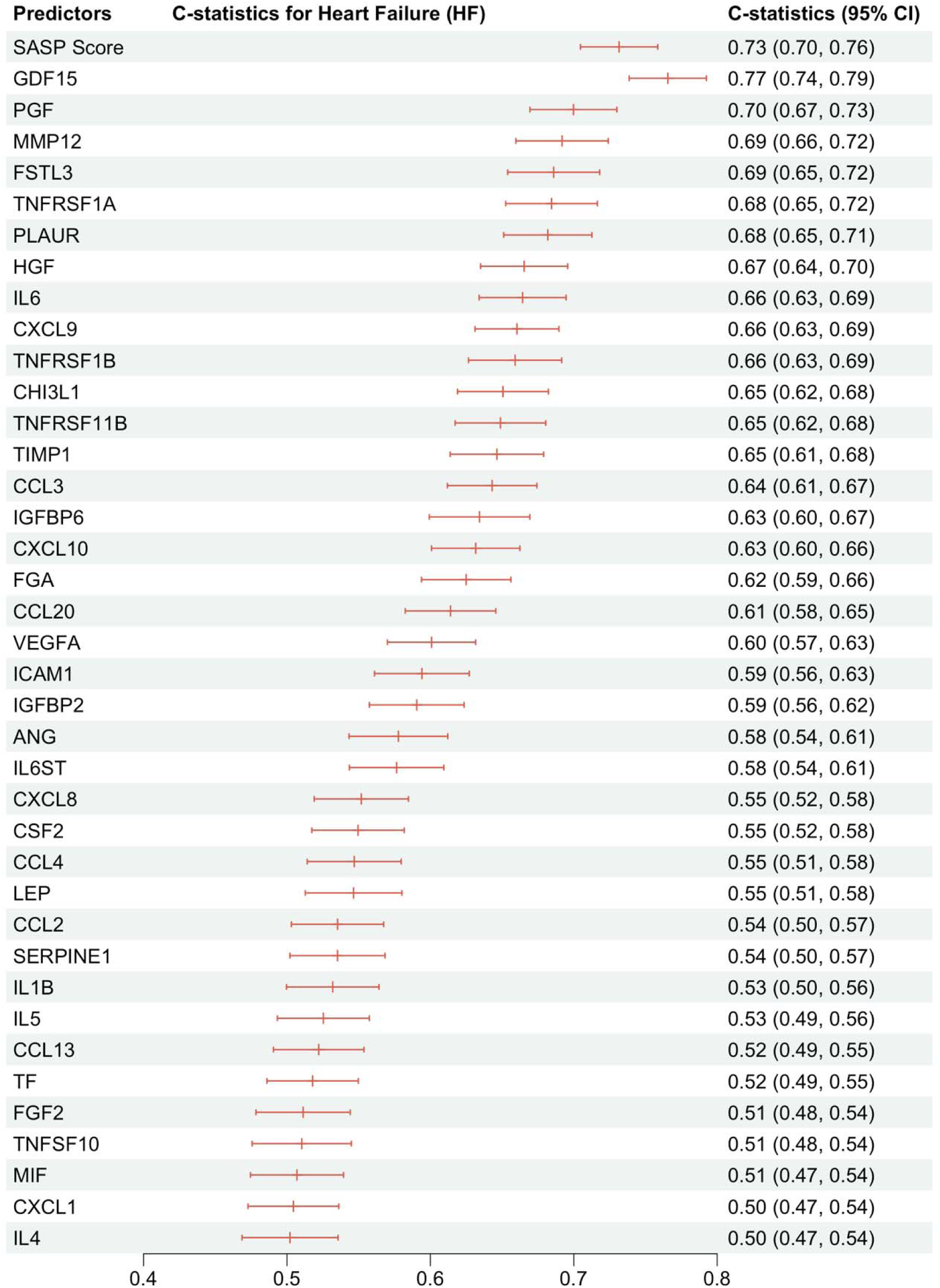
[UKB test samples (n=7,639)] C-statistics for incident heart failure predicted by baseline SASP score and individual proteins.

**Fig. S17.**
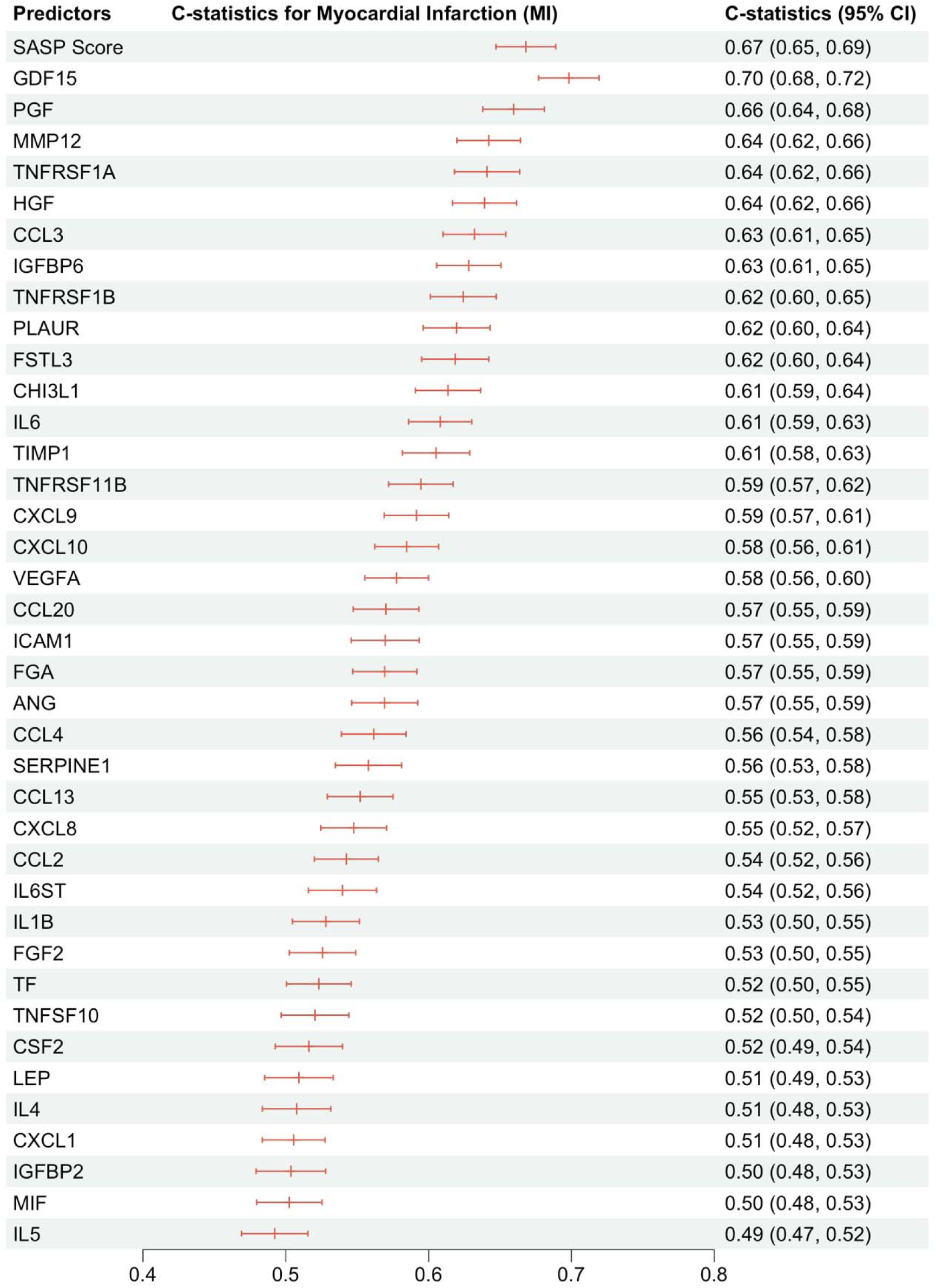
[UKB test samples (n=7,639)] C-statistics for incident myocardial infarction predicted by baseline SASP score and individual proteins.

**Fig. S18.**
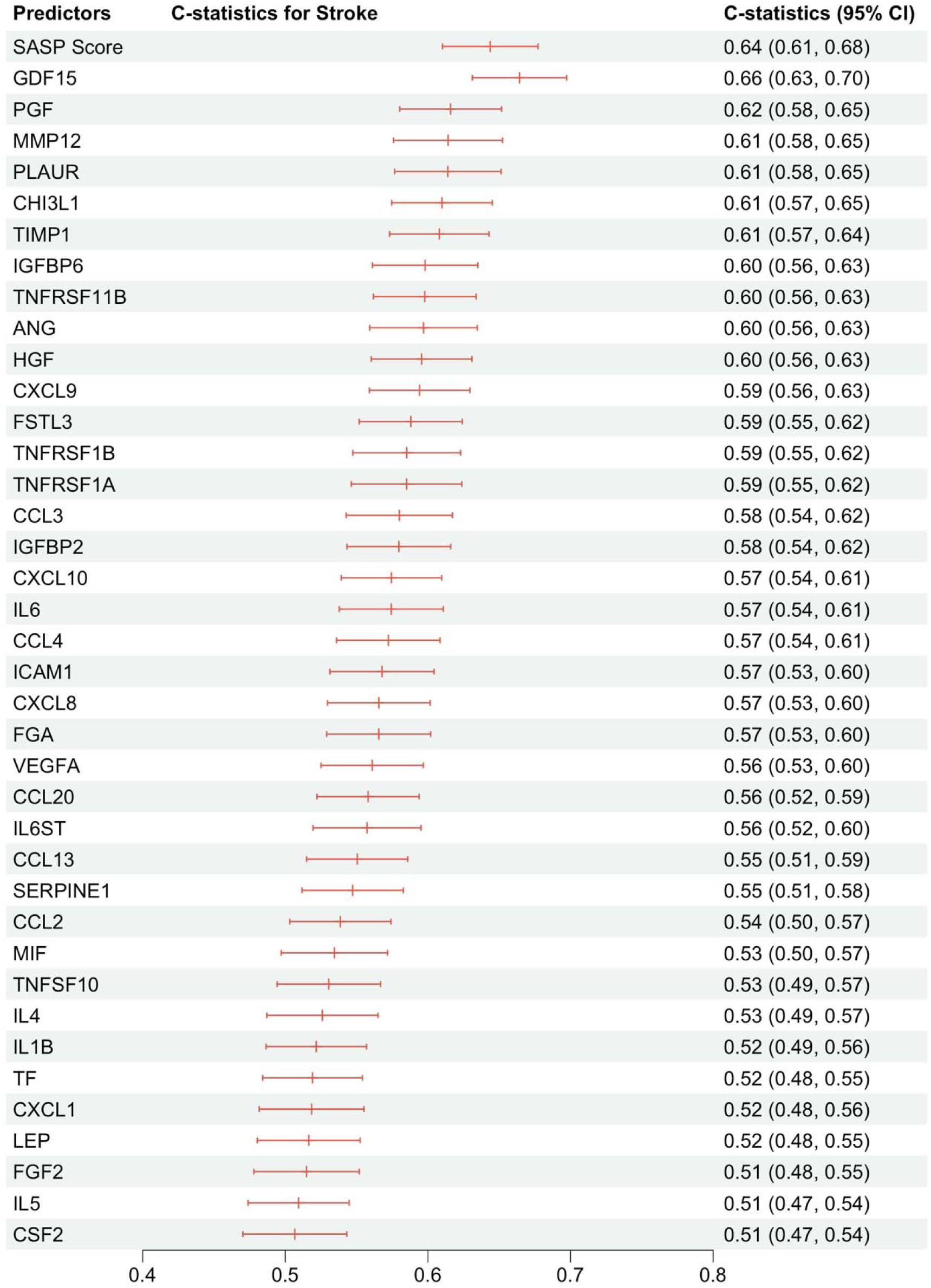
[UKB test samples (n=7,639)] C-statistics for incident stroke predicted by baseline SASP score and individual proteins.

**Fig. S19.**
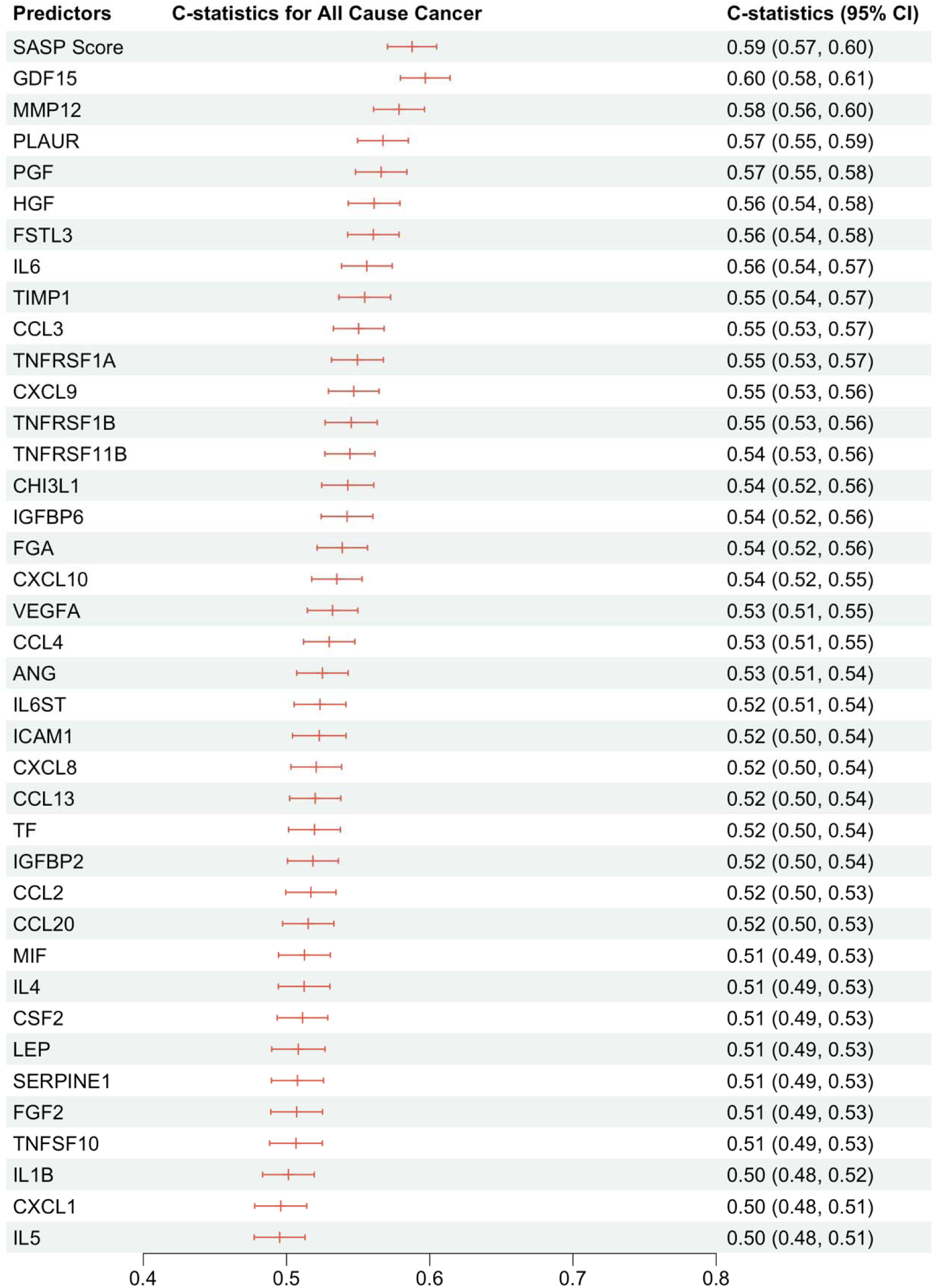
[UKB test samples (n=7,639)] C-statistics for incident all cause cancer predicted by baseline SASP score and individual proteins.

**Fig. S20.**
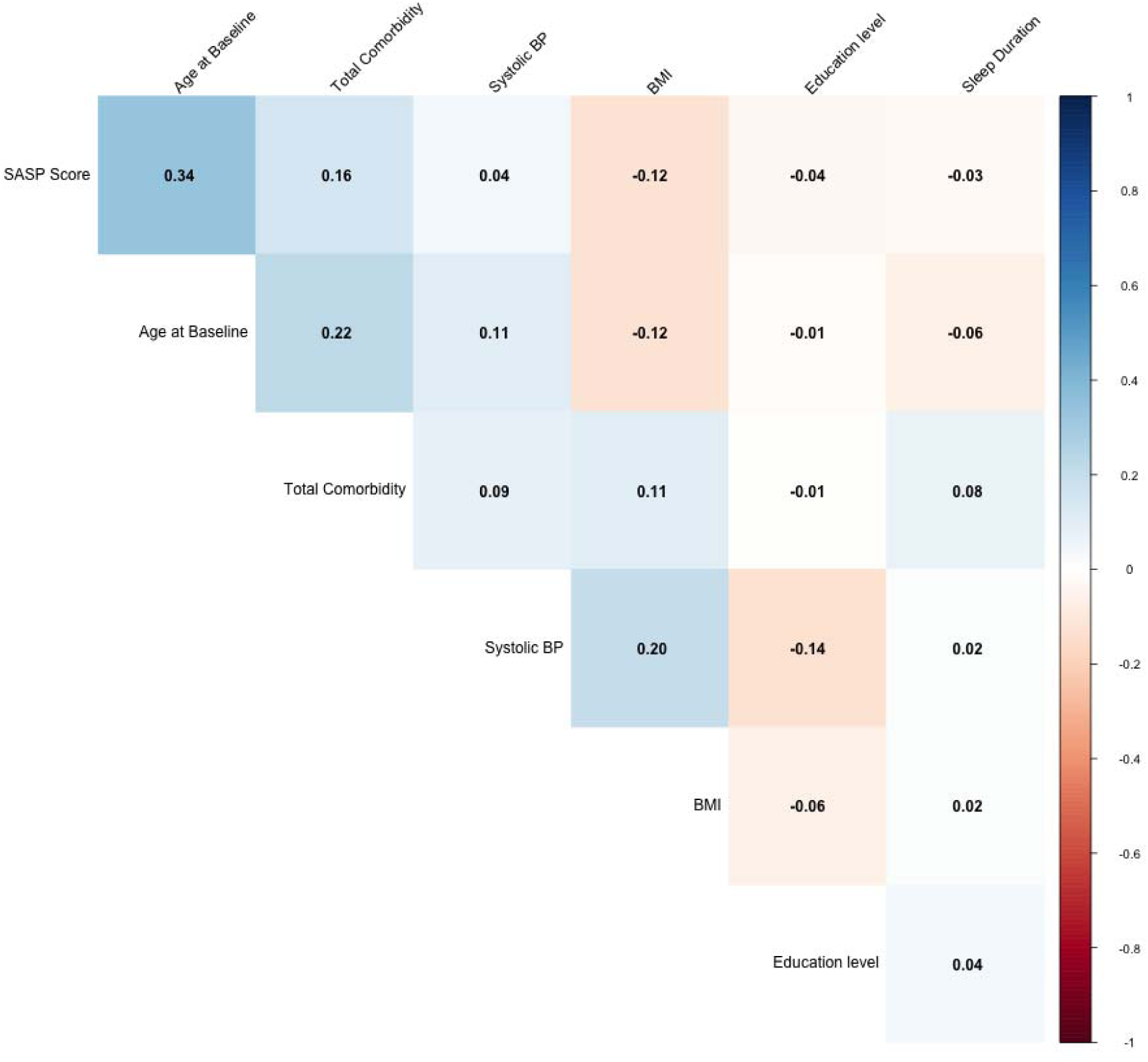
[MEDEX test samples (n=585)] Spearman’s correlation coefficients between SASP Score and baseline chronological age, BMI, Rockwood frailty, diastolic blood pressure, systolic blood pressure, leukocyte telomere length.

**Fig. S21.**
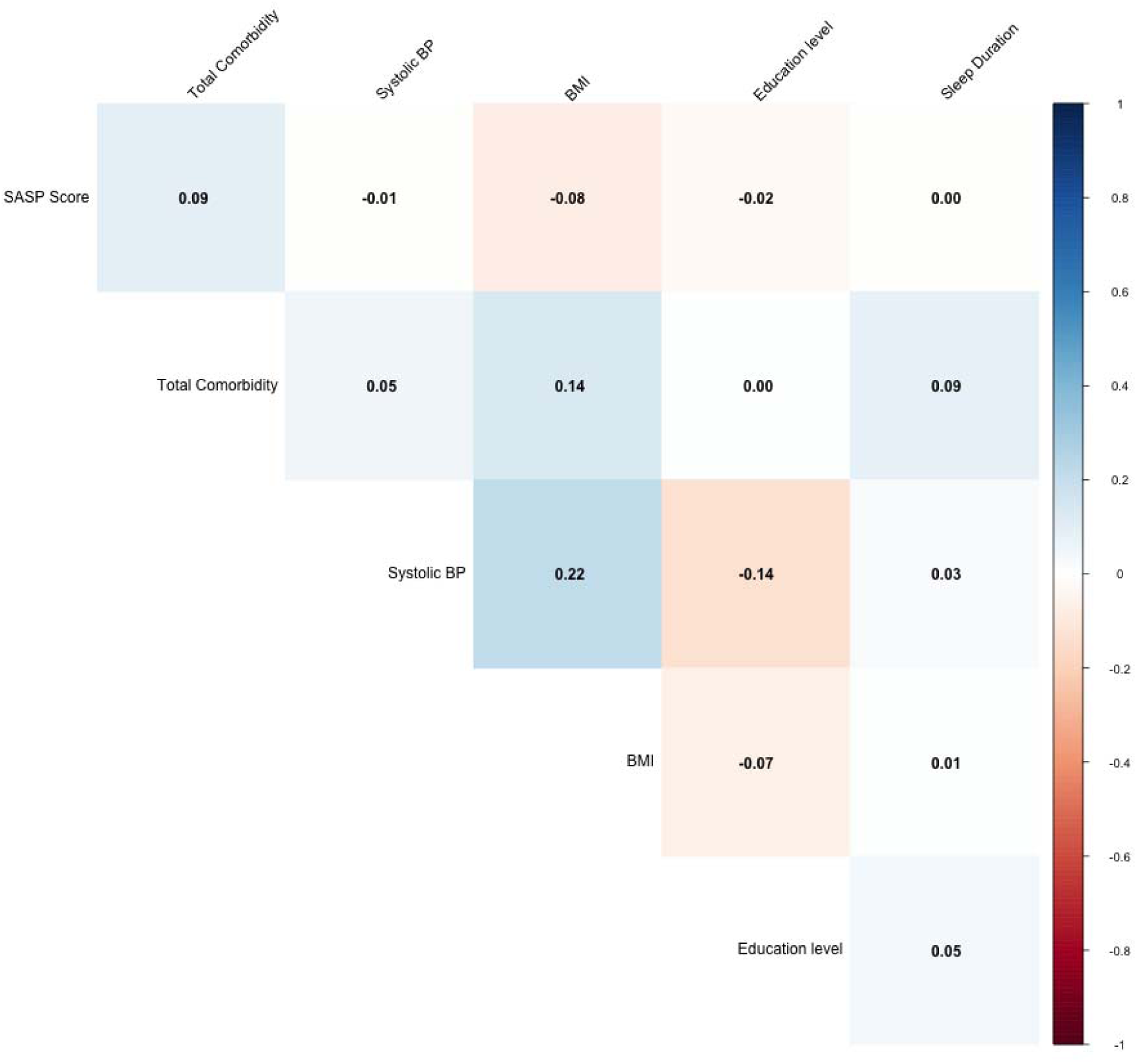
[MEDEX test samples (n=585)] Partial Spearman’s correlation coefficients between SASP Score and BMI, Rockwood frailty, diastolic blood pressure, systolic blood pressure, leukocyte telomere length, controlling for baseline chronological age.

**Fig. S22.**
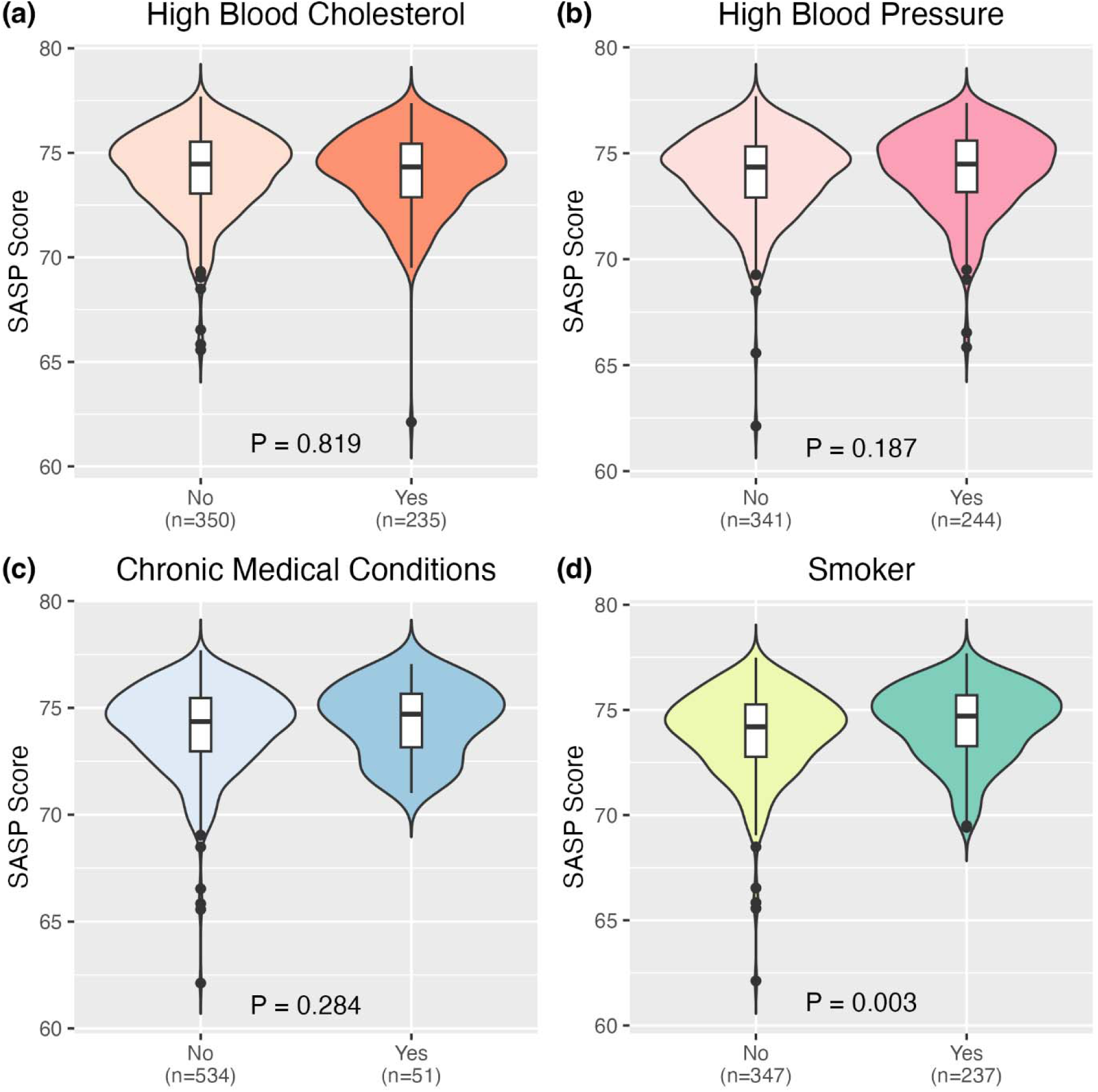
[MEDEX (n=585)] Baseline SASP Score distribution comparing people with or without different medical conditions at baseline (a) high blood cholesterol, (b) high blood cholesterol, (c) chronic medical conditions (diabetes, kidney diseases, heart attack, heart disease) (d) previous or current smoker.

## Supplementary Tables

**Table S1.**
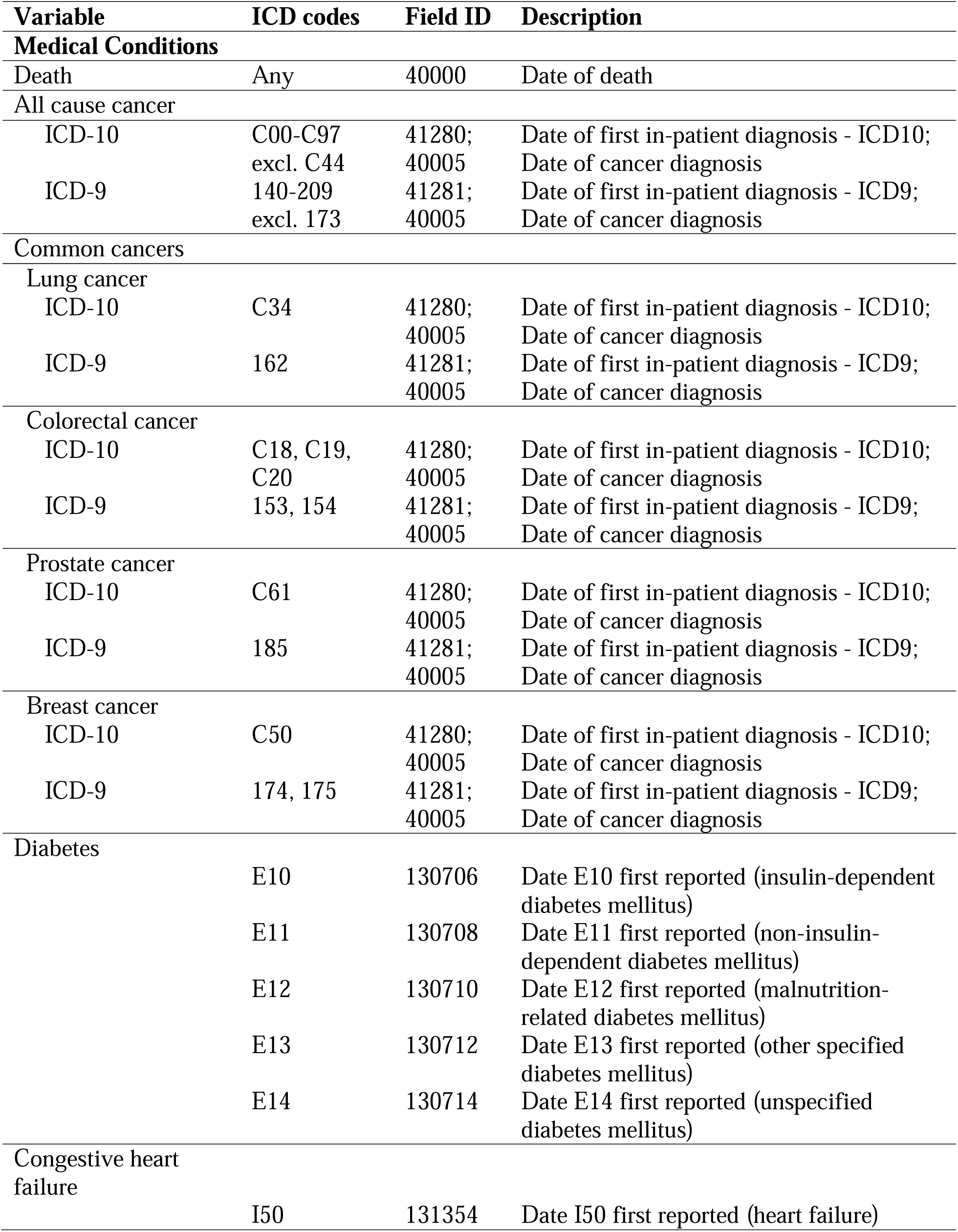
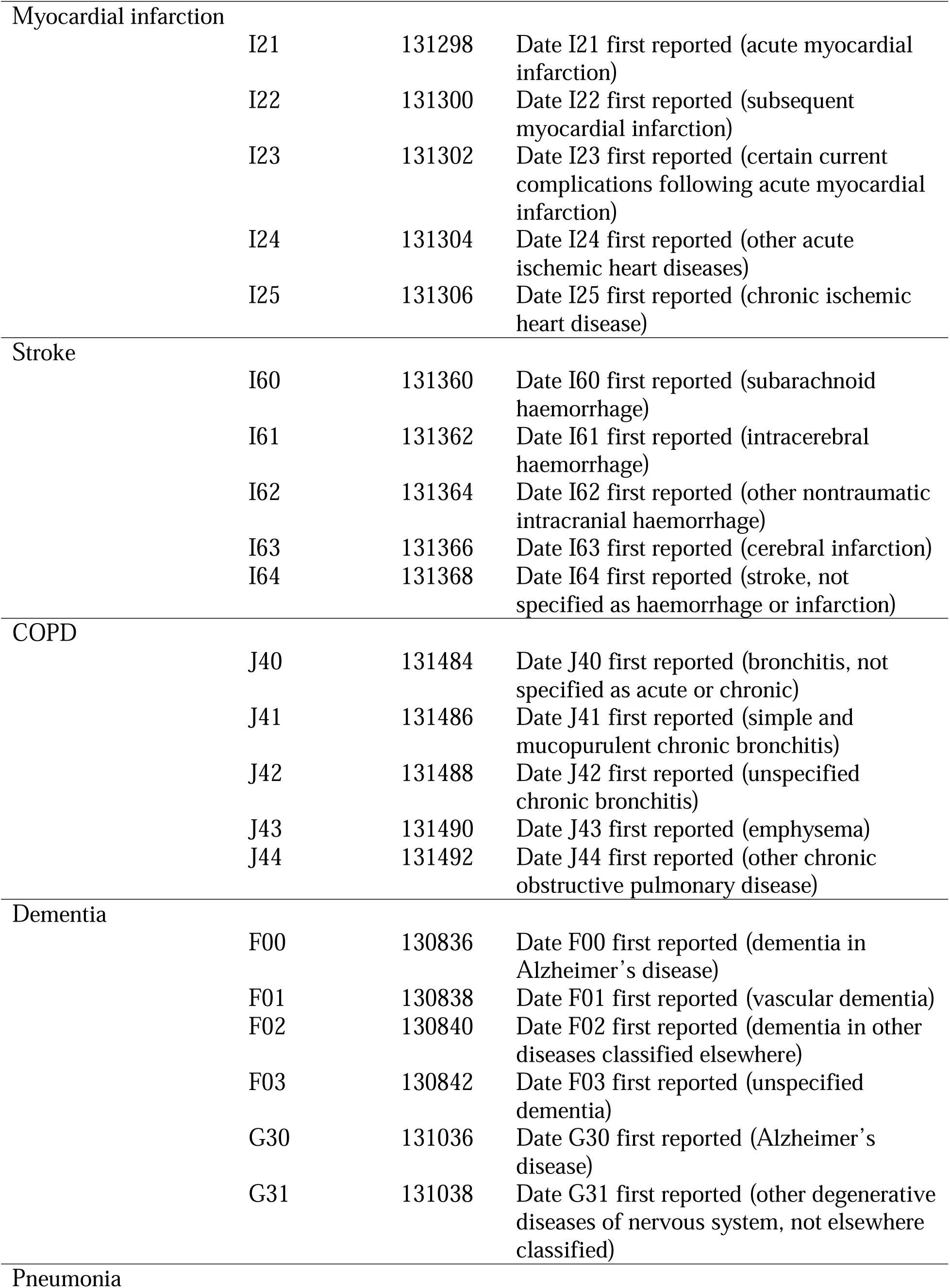
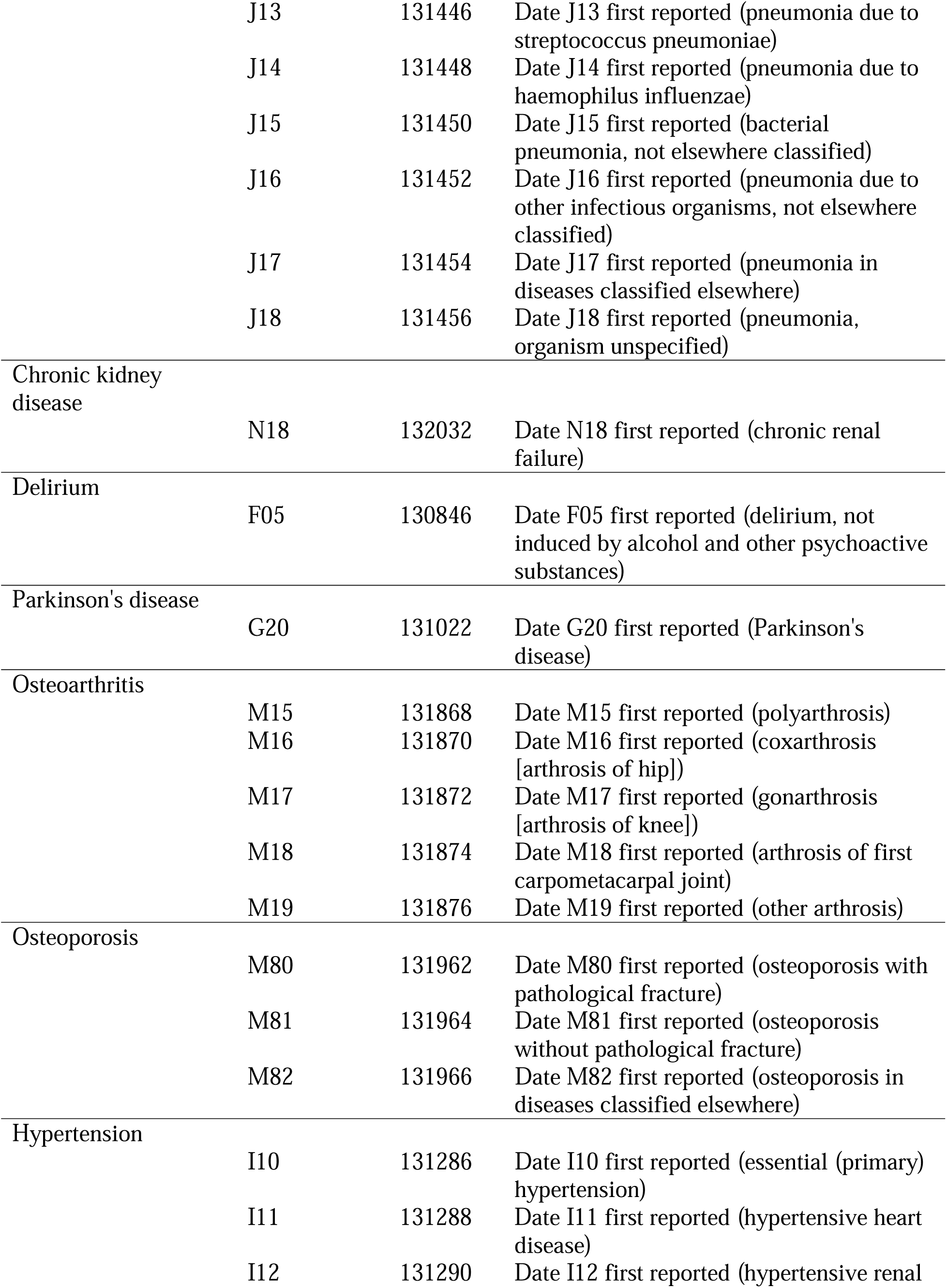
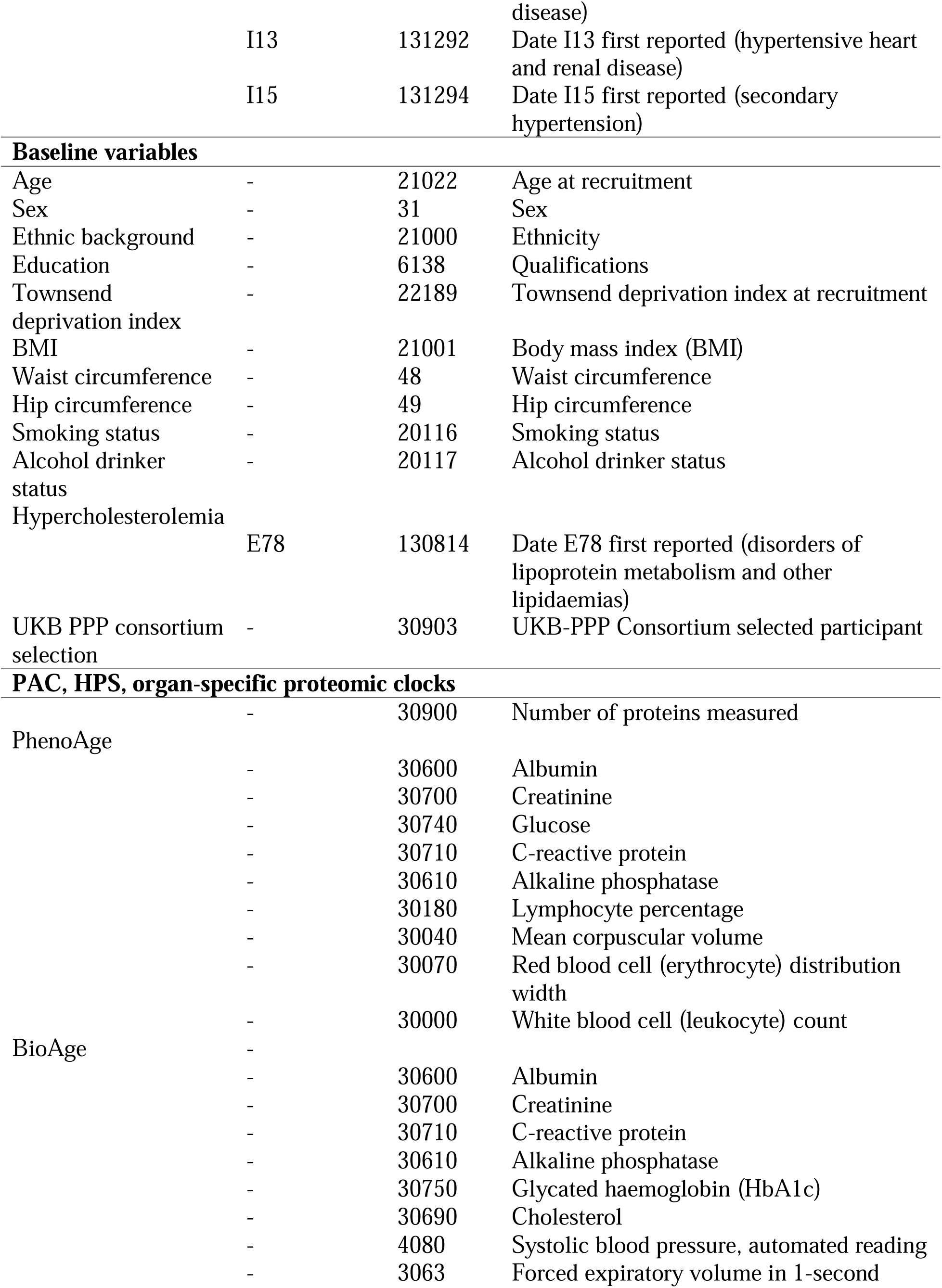
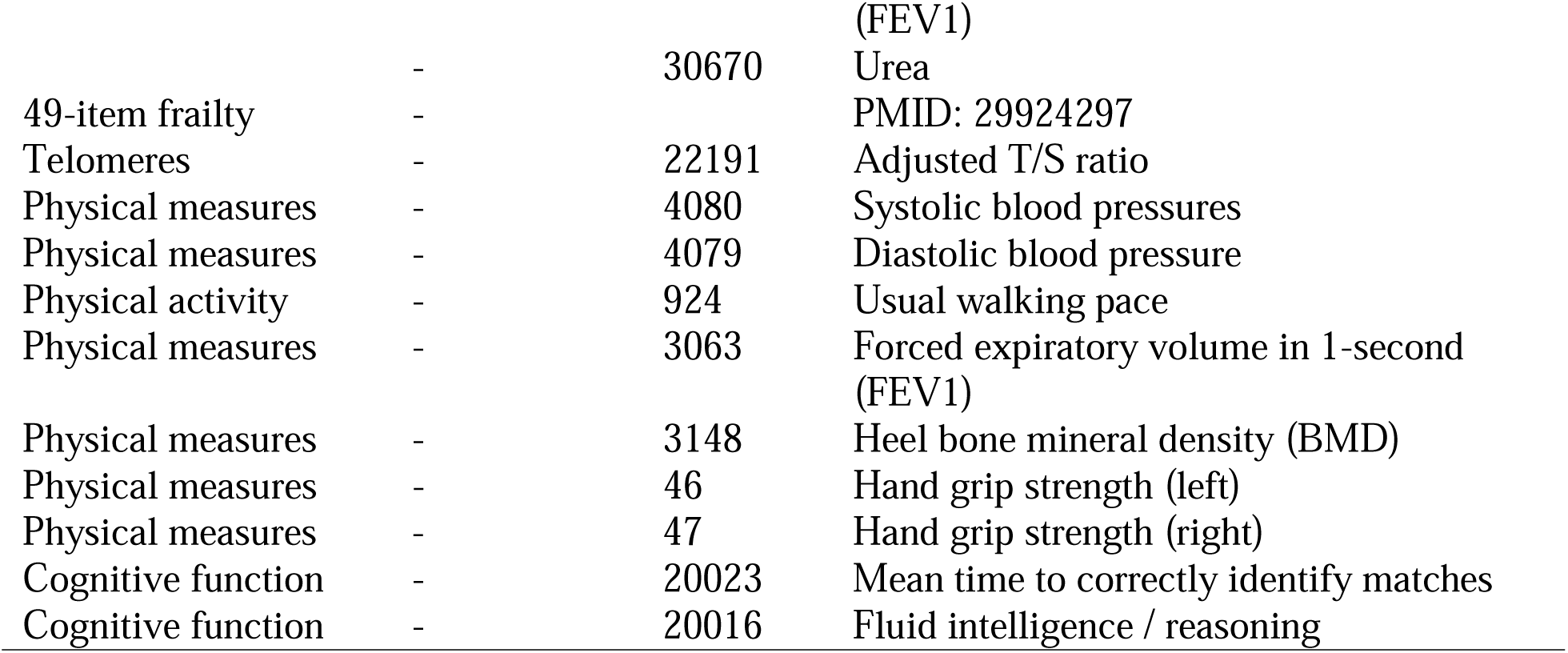
A list of UK Biobank field IDs to extract data for medical conditions, and baseline variables.

**Table S2.** UKB PPP participants (n=50,997) versus UKB baseline cohort participants not included in the UKB PPP (n=450,960)

| Baseline participant characteristics | UKB-PPP Participation |  |
| --- | --- | --- |
|  | No, N = 450,960<sup>I</sup> | Yes, N = 50,997<sup>I</sup> |
| Age | 56.50 ± 8.08; 58.00 (50.00, 63.00) | 56.82 ± 8.20; 58.00 (50.00, 64.00) |
| Sex |  |  |
| Male | 205,437 (45.56%) | 23,463 (46.02%) |
| Female | 245,523 (54.44%) | 27,516 (53.98%) |
| Ethnicity |  |  |
| White | 424,588 (94.64%) | 47,588 (93.75%) |
| Black | 7,814 (1.74%) | 1,277 (2.52%) |
| Asian | 11,091 (2.47%) | 1,178 (2.32%) |
| Other | 5,131 (1.14%) | 715 (1.41%) |
| Education |  |  |
| College or University degree | 144,581 (32.75%) | 16,350 (32.48%) |
| A levels/AS levels (or equivalent) | 49,639 (11.24%) | 5,602 (11.13%) |
| O levels/GCSEs (or equivalent) | 94,484 (21.40%) | 10,590 (21.04%) |
| CSEs (or equivalent) | 24,138 (5.47%) | 2,733 (5.43%) |
| NVQ or HND or HNC (or equivalent) | 29,339 (6.65%) | 3,366 (6.69%) |
| Other professional qualifications | 23,065 (5.22%) | 2,712 (5.39%) |
| None of the above | 76,232 (17.27%) | 8,989 (17.86%) |
| Townsend deprivation index | -1.31 ± 3.08; -2.14 (-3.65, 0.53) | -1.18 ± 3.18; -2.05 (-3.62, 0.76) |
| Drinking status |  |  |
| Never | 19,936 (4.44%) | 2,417 (4.75%) |
| Previous | 16,085 (3.58%) | 1,994 (3.92%) |
| Current | 413,421 (91.99%) | 46,434 (91.32%) |
| Smoking status |  |  |
| Never | 245,650 (54.80%) | 27,573 (54.35%) |
| Previous | 155,060 (34.59%) | 17,779 (35.04%) |
| Current | 47,547 (10.61%) | 5,382 (10.61%) |
| BMI | 27.43 ± 4.80; 26.74 (24.13, 29.91) | 27.47 ± 4.80; 26.78 (24.19, 29.92) |
| Waist to hip circumference ratio | 0.87 ± 0.09; 0.87 (0.80, 0.94) | 0.87 ± 0.09; 0.88 (0.80, 0.94) |
| Maximum grip strength | 32.65 ± 11.31; 30.00 (24.00, 40.00) | 32.54 ± 11.31; 30.00 (24.00, 40.00) |
| Hypertension | 120,248 (26.66%) | 14,372 (28.19%) |
| Hypercholesterolemia | 67,233 (14.91%) | 8,168 (16.02%) |

<sup>I</sup>Mean ± SD; Median (IQR) or Frequency (%)

**Table S3.**
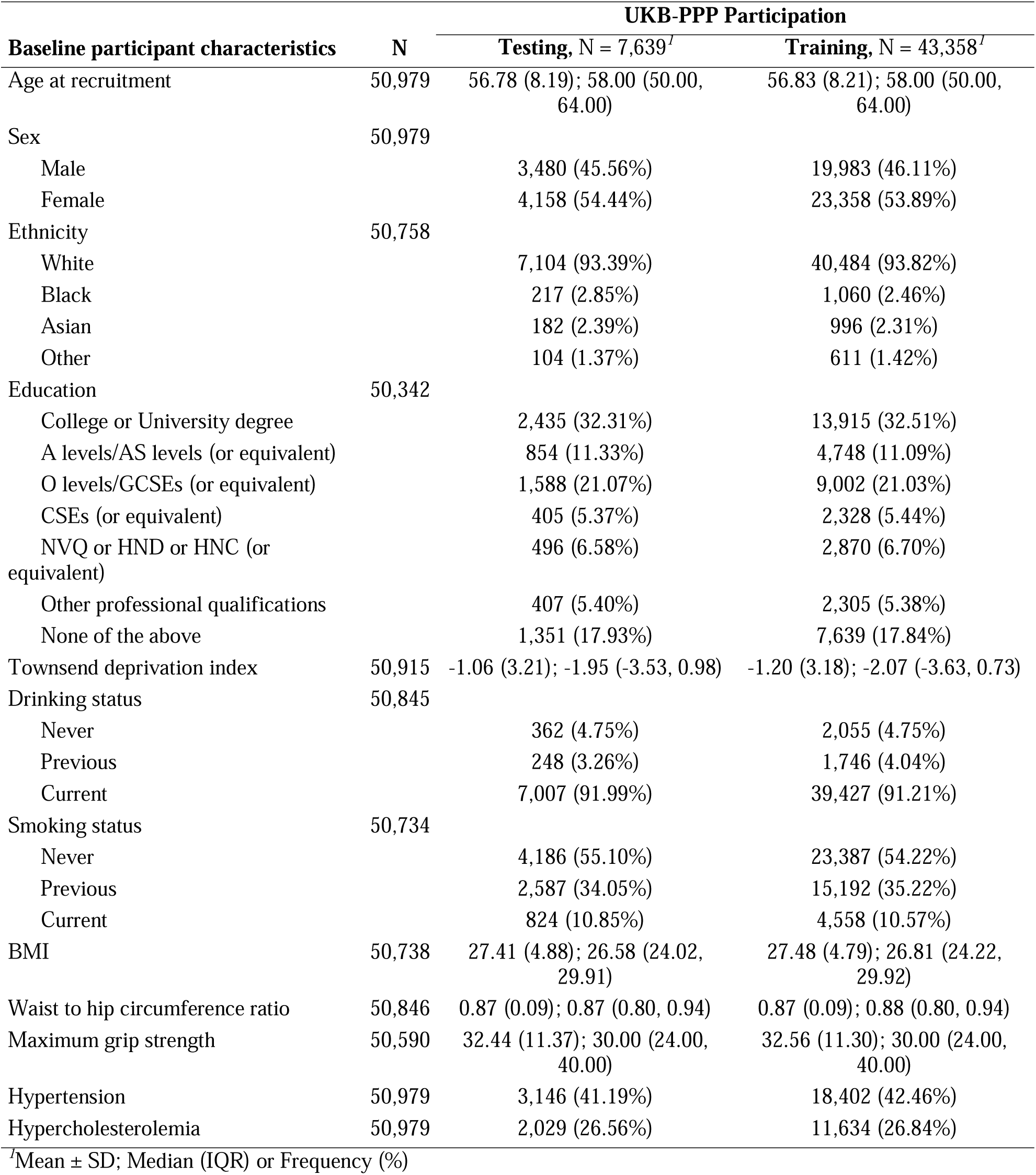
UKB PPP participants in the training (n=30,184) versus test set (n=12,935)

**Table S4.** MEDEX participants in the exercise group (n = 282) versus non-exercise group (n = 303)

| Participant characteristics | Exercise Group |  | Statistics | p-value<sup>2</sup> |
| --- | --- | --- | --- | --- |
|  | No, N = 303<sup>1</sup> | Yes, N = 282<sup>1</sup> |  |  |
| SASP Score at Baseline | 73.95 ± 1.94; 74.25 (72.73, 75.41) | 74.24 ± 1.95; 74.57 (73.21, 75.63) | 45770 | 0.042 |
| SASP Score at 6-month | 74.22 ± 1.90; 74.57 (73.14, 75.60) | 74.34 ± 1.98; 74.70 (73.35, 75.70) | 40172 | 0.295 |
| SASP Score at 18-month | 74.37 ± 1.77; 74.64 (73.61, 75.73) | 74.39 ± 2.00; 74.72 (73.62, 75.71) | 26860 | 0.697 |
| Age | 71.18 ± 4.43; 70.00 (68.00, 74.00) | 71.80 ± 5.17; 70.50 (67.00, 75.00) | 44610 | 0.355 |
| Sex |  |  |  | 0.461 |
| Female | 215 (70.96%) | 208 (73.76%) |  |  |
| Male | 88 (29.04%) | 74 (26.24%) |  |  |
| Race |  |  |  | 0.007 |
| White | 256 (84.49%) | 221 (78.37%) |  |  |
| Black or African American | 37 (12.21%) | 32 (11.35%) |  |  |
| Asian | 8 (2.64%) | 19 (6.74%) |  |  |
| Others | 2 (0.66%) | 10 (3.55%) |  |  |
| Ethnicity |  |  |  | 0.186 |
| Hispanic or Latino | 16 (5.28%) | 23 (8.16%) |  |  |
| NOT Hispanic or Latino | 287 (94.72%) | 259 (91.84%) |  |  |
| Education Level | 16.04 ± 2.13; 16.00 (14.00, 18.00) | 16.29 ± 2.25; 16.00 (14.00, 18.00) | 45501 | 0.158 |
| Total Comorbidity | 6.74 ± 2.97; 7.00 (5.00, 9.00) | 6.82 ± 2.81; 7.00 (5.00, 9.00) | 43300 | 0.776 |
| Systolic Blood Pressure at Baseline | 135.42 ± 15.95; 134.00 (126.00, 147.00) | 136.41 ± 16.81; 135.00 (125.00, 146.00) | 43106 | 0.687 |
| Diastolic Blood Pressure at baseline | 76.25 ± 10.55; 77.00 (69.00, 83.00) | 77.20 ± 10.48; 78.00 (71.00, 85.00) | 44772 | 0.221 |
| BMI at Baseline | 28.08 ± 5.24; 27.00 (24.00, 31.00) | 28.66 ± 4.97; 28.00 (25.00, 32.00) | 46210 | 0.087 |

<sup>1</sup>Mean ± SD; Median (IQR) or Frequency (%)
<sup>2</sup>Wilcoxon rank sum test; Fisher's exact test

