## Supplementary Methods for "A deep-learning based biomarker of systemic cellular senescence burden to predict mortality and health outcomes"

**Guided Auto-Encoder with Transformers (GAET) and the Development of SASP Scores**

We aimed to develop a model for extreme dimensionality reduction, while preserving complex, nonlinear interactions within a protein network. The resulting output scalar score could be directly used in downstream applications as a proxy for biological process. To do so, we introduced the Guided Auto-Encoder with Transformers (GAET). This model is built on the foundations of the autoencoder and the guided autoencoder framework. A classic guided autoencoder^1^ is designed to generate informative representation towards a specific target. However, conventional guided autoencoders use basic multilayer perceptron for core model structure, limiting its ability to model and memorize complex relationships across protein interaction networks. We therefore proposed the Guided Auto-Encoder with Transformers, where the Transformer^2^ module functioning as the core for both encoder and decoder. This new design substantially enhances the model’s capacity to capture complex network interaction patterns, making it possible to translate models and borrow structural information cross cohorts. We applied GAET to derive SASP Scores. It’s a nonlinear summarization of SASP data, portraying a latent factor of cellular senescence.

**Guided Auto-Encoder**

The guided autoencoder is built based on classic autoencoder, which is designed for unsupervised dimension reduction. Typical autoencoders consist of two components, an encoder to compress the input data, and a decoder to reconstruct input data from the compressed data. Given any input data $x$, autoencoders were trained to minimize the reconstruction loss

$$\mathcal{L}_{recon}=\left( f_{AE}\left( x,\theta\right)-x \right)^{2},$$

where we denote $f_{AE}(x,\theta)$ as the output from autoencoder with parameters set $\theta$. By doing so, the network is supposed to learn a compressed representation of the original data that best reconstruct it.

Guided autoencoder is designed with two focuses: (1) the learned compact representation can be recovered from the original data as much as possible (reconstruction loss) and (2) the learned compact representation should be as informative of the desired target as possible (guidance loss). Given the input $x$, a phenotype or biomarker $y$ as guidance, the guidance loss is defined as

$$\mathcal{L}_{pred}=\left( f_{G}\left( x,\theta\right)-y \right)^{2},$$

where $f_{G}(x,\theta)$ is a 1-D linear combination of the model bottleneck, which would be the SASP Score to train. A GAE aims to reduce both reconstruction loss and guidance loss

$$\mathcal{L=}{\lambda\mathcal{L}}_{recon}+\left( 1-\lambda\right)\mathcal{L}_{pred},$$

where $\lambda$ is a tunable parameter to balance the two terms. The $\lambda$ was determined when the empirical compound loss is minimized. In practice, pointwise mean squared error (MSE) is used for both loss terms. Chronological age was chosen as the guidance, giving its widespread availability, thereby allowing this model to be translated cross multiple cohorts.

**Transformer**

Transformer is a neural network architecture based on a multi-head attention mechanism. Originally developed for natural language processing, it has been adapted to protein informatics^3^ for two key reasons. First, its ability to model long-range dependencies and maintain content awareness makes it well-suited to capture and decode the complex relationships among proteins. Second, its capacity to handle variable-length inputs allows the model to be trained and applied directly to data with missing values or datasets with different distributions, potentially eliminating the need for extensive imputation and enabling applicability across studies using different proteomics analysis platforms. These properties position the Transformer as a natural choice for our proteomics autoencoder. To combine the semantic information of proteins, the Transformer decoder takes in not only the real valued proteomic data, but also numeric labels representing the names of each protein. Note that the data flow’s dimensionality remains changed after processed by the Transformer encoder and decoder module.

**Guided Autoencoder with Transformer**

We introduce the Guided Autoencoder (GAET), which comprises three components: (i) a Transformer encoder, (ii) a Transformer decoder, and (iii) a linear guidance head.

The model consumes a proteomics data matrix $X\in\mathbb{R}^{N\times P}$, where $N$ is the number of samples and $P$ is the number of proteins. A length-$P$ vector of protein identifiers $\left( t_{1},\ldots,t_{P} \right)$ aligned to the columns of $X$ is also needed for corresponding learned protein embeddings. Let $x_{i}\in\mathbb{R}^{P}$denote the measurements for sample $i$. Each protein index $t_{j}$ is mapped to a learnable protein token embedding $e_{j}\in\mathbb{R}^{D}$. The corresponding scalar measurement $x_{i,j}$ is projected to the model width embedding $s_{i,j}\in\mathbb{R}^{D}$. Because proteins are entities instead of ordered sequences, we did not add positional encodings. The input token for protein $j$ in sample $i$ is the sum

$$u_{i,j}=e_{j}+s_{i,j}\in\mathbb{R}^{D},$$

yielding a sequence $U_{i}=[u_{i,1},\ldots,u_{i,P}]\in\mathbb{R}^{P\times D}$ for each sample. The sequences are then fed to a Transformer encoder, which returns $H\in\mathbb{R}^{N\times P\times D}$, where $H_{i,j,:}$ is a contextual embedding for protein $j$ in sample $i$, integrating information from all observed proteins. A sample-level summarization is then obtained by pooling over the sequence dimension, aggregating each sample’s token embeddings to produce an $N\times D$ representation. A final fully connected layer projects the pooled representation to a latent space, yielding the bottleneck $Q\in\mathbb{R}^{N\times L}$, where $L<P\ll D$.

From the encoder’s latent representation, the network forks into two branches. The first branch starts with a linear layer that expands $Q$ to the model width $D$, producing $H^{0}\in\mathbb{R}^{N\times D}$. To accommodate variable sequence length $P$ at training and inference, $H^{0}$ is repeatedly broadcasted along the sequence axis to form the decoder target $\tilde{H}\in\mathbb{R}^{N\times P\times D}$ of each sample. The decoder output is then passed through a token-wise linear layer to produce the reconstructed proteomics data matrix $\hat{X}\in\mathbb{R}^{N\times P}$.

The other branch from the encoder output is a fully connected linear guidance layer that maps $Q$ to a single scalar $y_{i}\in\mathbb{R}$ per sample. In our implementation, this guidance head predicts chronological age and is trained jointly with reconstruction. The resulting $y_{i}$ is the SASP Score for each sample.
